# Early versus Delayed Initiation of Renal Replacement Therapy in Critically Ill Patients with Acute Kidney Injury: A Systematic Review and Meta-Analysis of Randomized Trials

**DOI:** 10.1101/2025.09.17.25336038

**Authors:** Rakhshanda Khan, Sri Mani Krishna Satvik Matta, Gayathri Boda, Madhamshetty Shivani, Tetta Venkata Sarma Jayant, Shankar Biswas, Elavarasi Kuppan, Yasaswini Nimmagadda, Amatul Aziz, Harshawardhan Dhanraj Ramteke

## Abstract

**Introduction:** Acute kidney injury (AKI) affects up to 66% of critically ill patients and is associated with substantial morbidity and mortality. Renal replacement therapy (RRT) remains the cornerstone of management in severe AKI, yet the optimal timing of initiation remains controversial. While early RRT may rapidly correct metabolic derangements, it may expose patients to unnecessary risks. We performed an updated systematic review and meta-analysis to evaluate the effects of early versus delayed RRT initiation on mortality and adverse outcomes in critically ill patients with AKI.

**Methods:** We systematically searched PubMed, Embase, Cochrane CENTRAL, and Web of Science up to August 2025 for randomized controlled trials (RCTs) comparing early versus delayed RRT initiation in adult ICU patients with AKI. Two reviewers independently screened studies, extracted data, and assessed risk of bias using RoB 2.0. Pooled analyses were conducted using a random-effects model. Subgroup analyses were performed by age (<65 vs ≥65 years) and illness severity (SOFA <12 vs ≥12). Quality of evidence was graded using GRADE.

**Results:** A total of **19 RCTs comprising 12,162 patients** were included. Of these, **5,450 patients were randomized to early RRT** and **5,452 to delayed RRT**. The mean initiation time was **10.5 hours in early groups** versus **35.3 hours in delayed groups**. Overall, early initiation **did not significantly reduce 30-day mortality** (RR = 0.96, 95% CI: 0.89–1.04; p = 0.31) or **90-day mortality** (RR = 0.90, 95% CI: 0.76–1.06; p = 0.17). Subgroup analyses showed no survival benefit across age (<65 vs ≥65) or SOFA (<12 vs ≥12) strata. Adverse events analysis revealed that early initiation was associated with a **higher risk of infection** (RR = 1.21, 95% CI: 1.03–1.39; p = 0.02), while no significant differences were found in **hypotension** (RR = 1.03, 95% CI: 0.81–1.25), **arrhythmias** (RR = 1.10, 95% CI: 0.78–1.42), or **bleeding events** (RR = 0.93, 95% CI: 0.75–1.11).

**Conclusion:** Early initiation of RRT in critically ill patients with AKI **does not improve short- or long-term mortality** and is associated with an **increased risk of infection**. These findings support a **delayed strategy**, reserving RRT for patients with absolute indications or those not improving with conservative management. Individualized decision-making based on clinical status and illness trajectory should remain the standard of care.

## Introduction

Acute kidney injury (AKI) is one of the most frequent complications in critically ill patients, affecting up to 70% of those admitted to intensive care units (ICUs) [1]. Severe AKI is associated with hospital mortality rates as high as 50–60% [2]. The abrupt decline in renal function leads to life-threatening complications including fluid overload, electrolyte imbalance, metabolic acidosis, and accumulation of uremic toxins [3].

Renal replacement therapy (RRT) remains the cornerstone of management when such derangements occur. Guidelines such as those from KDIGO recommend RRT in the presence of refractory hyperkalemia, severe metabolic acidosis, or pulmonary edema [4]. RRT rapidly corrects these abnormalities, thereby stabilizing patients [5]. However, the optimal timing of RRT initiation in patients without urgent indications is still debated. Early initiation may prevent further complications and reduce mortality [6], but it may also expose patients— whose renal function could recover spontaneously—to risks such as intradialytic hypotension, bleeding, infection, and vascular access–related complications [7].

Several randomized controlled trials (RCTs) have addressed this controversy. The ELAIN trial suggested a mortality benefit with early initiation [8], while the AKIKI [9] and IDEAL-ICU [9] trials did not confirm such an effect. More recently, the large multicenter STARRT-AKI trial reported no mortality benefit with accelerated RRT compared to a standard strategy [10]. These conflicting findings have been mirrored in prior meta-analyses, which reached inconsistent conclusions due to heterogeneity and methodological limitations [11]. A recent updated meta-analysis [12] and a 2023 Cochrane Review [13] continue to highlight the uncertainty, despite incorporation of newer large-scale trials.

In light of these developments, we conducted an updated systematic review and meta-analysis of RCTs comparing early versus delayed RRT initiation in critically ill patients with AKI. Our objectives were to evaluate the impact of timing on mortality, renal recovery, RRT duration, and adverse events. We also assessed the certainty of evidence using the Grading of Recommendations, Assessment, Development, and Evaluation (GRADE) framework.

## Methods

### Literature Search

We conducted a comprehensive literature search in PubMed, Embase, Cochrane Central Register of Controlled Trials (CENTRAL), and Web of Science from inception to September 2025. The search strategy combined controlled vocabulary and free-text terms related to acute kidney injury, renal replacement therapy, dialysis, timing, early initiation, and delayed initiation. Reference lists of relevant articles and prior systematic reviews were also screened. We limited the search to randomized controlled trials involving critically ill adult patients. No language restrictions were applied. The full search strategy for each database is provided in the supplementary material. Prisma protocol was followed and registered with Prospero with number CRD420251150278 [14].

### Study Selection and Data Extraction

Two reviewers independently screened all retrieved titles and abstracts to identify potentially eligible studies. Full texts of relevant articles were assessed according to predefined inclusion and exclusion criteria. Only randomized controlled trials (RCTs) comparing early versus delayed initiation of renal replacement therapy (RRT) in critically ill adult patients with acute kidney injury (AKI) were included. Disagreements were resolved by discussion or by consulting a third reviewer.

For each eligible study, data were independently extracted using a standardized form. Extracted information included study characteristics (author, year, country, design, sample size), patient demographics, definitions of early and delayed RRT initiation, intervention details, modality of RRT, follow-up duration, and clinical outcomes (mortality, renal recovery, adverse events).

### Statistical Analysis

We performed the meta-analysis using Review Manager (RevMan, Cochrane Collaboration) and Stata software. For dichotomous outcomes (e.g., mortality, renal recovery, adverse events), we calculated risk ratios (RRs) with 95% confidence intervals (CIs). For continuous outcomes (e.g., duration of RRT, ICU stay), we extracted or calculated mean differences (MDs) or standardized mean differences (SMDs) with 95% CIs. A random-effects model (DerSimonian and Laird method) was applied to account for inter-study heterogeneity.

Statistical heterogeneity was quantified using the I² statistic, with values of 25%, 50%, and 75% indicating low, moderate, and high heterogeneity, respectively. Sensitivity analyses were conducted by sequentially excluding individual studies. We performed predefined subgroup analyses according to study design, patient population (septic vs. non-septic), and RRT modality. Potential publication bias was assessed visually using funnel plots and quantitatively using Egger’s regression test. The certainty of evidence was rated using the Grading of Recommendations, Assessment, Development, and Evaluation (GRADE) framework.

### Risk of Bias Assessment and Certainty of Evidence

The risk of bias for each included randomized controlled trial (RCT) was independently assessed by two reviewers using the Cochrane Risk of Bias 2.0 tool. This framework evaluates bias across five domains: (1) randomization process, (2) deviations from intended interventions, (3) missing outcome data, (4) measurement of the outcome, and (5) selection of the reported result. Each domain was graded as low risk, some concerns, or high risk. Discrepancies were resolved through consensus or adjudication by a third reviewer.

The overall certainty of evidence for each outcome was evaluated using the Grading of Recommendations, Assessment, Development, and Evaluation (GRADE) approach. Evidence was rated across the domains of study limitations (risk of bias), inconsistency, indirectness, imprecision, and publication bias. The quality of evidence was categorized as high, moderate, low, or very low. Summary of Findings (SoF) tables were generated to present the pooled effect estimates along with the corresponding GRADE ratings for primary and secondary outcomes.

## Result

### Demographics

A total of 315 studies were analyzed out of which 20 were selected [15–34] Figure 1. A total of 12,162 critically ill patients with acute kidney injury (AKI) were included across the eligible randomized controlled trials. Of these, 7,054 (58.0%) were male and 4,951 (40.7%) were female. The predominant modality used was continuous renal replacement therapy (CRRT), reported in the majority of studies, with smaller proportions receiving continuous venovenous hemofiltration (CVVH) and intermittent hemodialysis (IHD). Patients randomized to early RRT initiation began treatment at a mean of 10.5 hours, while those in the delayed group-initiated therapy at a mean of 35.3 hours. The overall mean SOFA score was 12.35 (SD 2.98), reflecting a severely ill cohort. Treatment groups were evenly balanced, with 5,450 patients in the early group and 5,452 in the delayed group.

**Figure 1.**
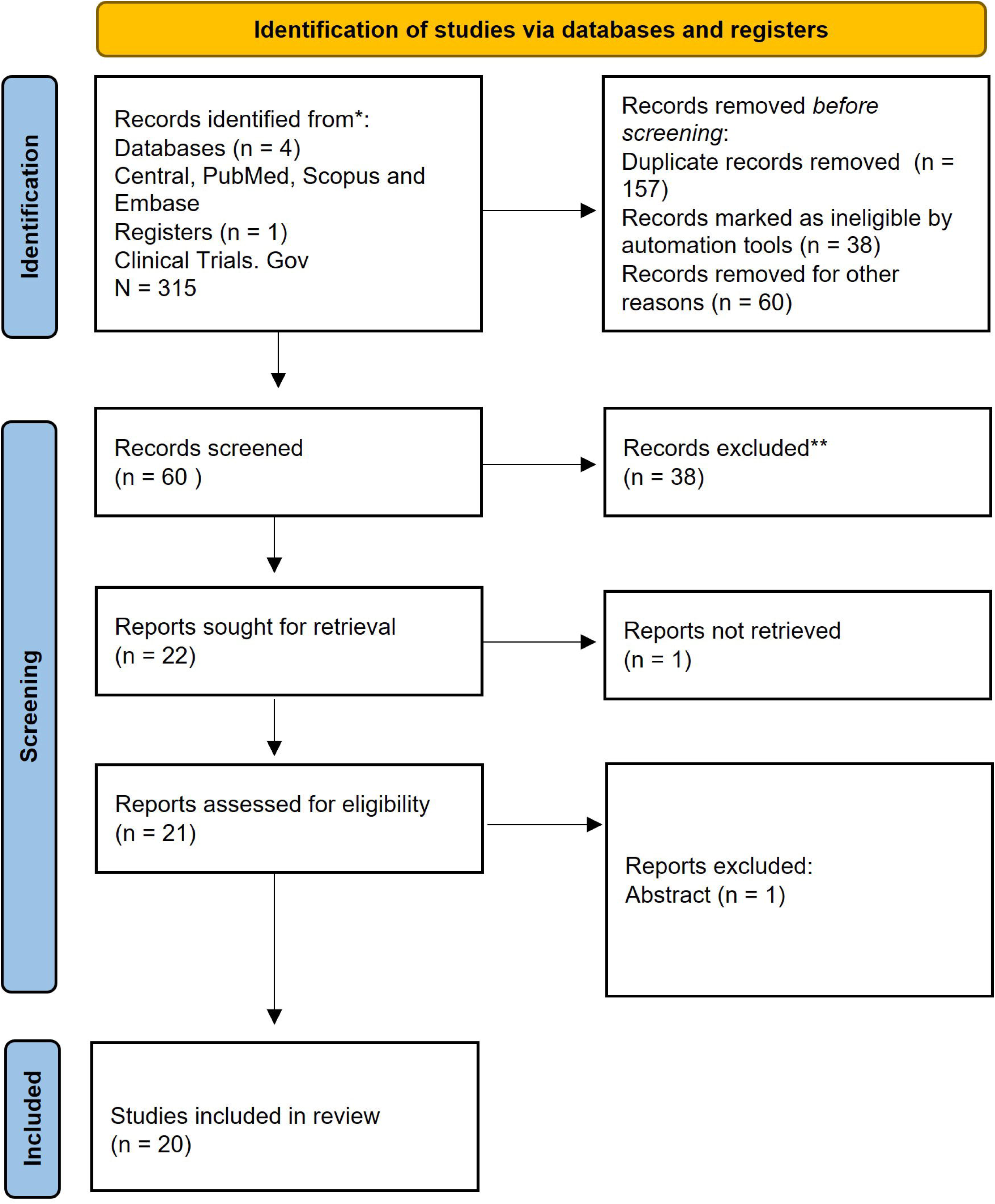
PRISMA Flow Diagram.

### Subgroup Analysis of 30-Day Mortality by SOFA Score

When stratified by baseline SOFA score, no significant difference in 30-day mortality was observed between early and delayed RRT initiation. In the SOFA <12 subgroup, pooled results from nine RCTs, including Bagshaw et al. (2020), Combes et al. (2015), and Lumlergul et al. (2018), showed no mortality benefit for early initiation (RR = 0.93, 95% CI: 0.73–1.19; p = 0.60), though heterogeneity was moderate (I² = 76%). Individual trials demonstrated variable effects: while Jamale et al. (2013) and Jiao et al. (2025) reported trends toward higher mortality with early initiation, Jun et al. (2014) suggested a possible protective effect with delayed therapy.

Similarly, in the SOFA >12 subgroup, seven RCTs—including STARRT-AKI (Bagshaw et al. 2020 subset), IDEAL-ICU (Barbar et al. 2018), and AKIKI-2 (Gaudry et al. 2021)—showed no survival advantage for early initiation (RR = 1.00, 95% CI: 0.81–1.19; p = 1.00), with high heterogeneity (I² = 82%). Again, individual trials varied: An et al. (2021) suggested reduced mortality with early initiation, whereas Jeong et al. (2025) reported higher mortality in the early group.

The overall pooled analysis across both strata confirmed the absence of mortality benefit (RR = 0.96, 95% CI: 0.81–1.11; p = 0.60), with no evidence of effect modification by SOFA score (test for subgroup differences, p = 0.68). These findings highlight the inconsistency across individual trials and reinforce that baseline illness severity, as measured by SOFA, does not appear to influence the impact of RRT timing on short-term survival. Figure 2.

**Figure 2.**
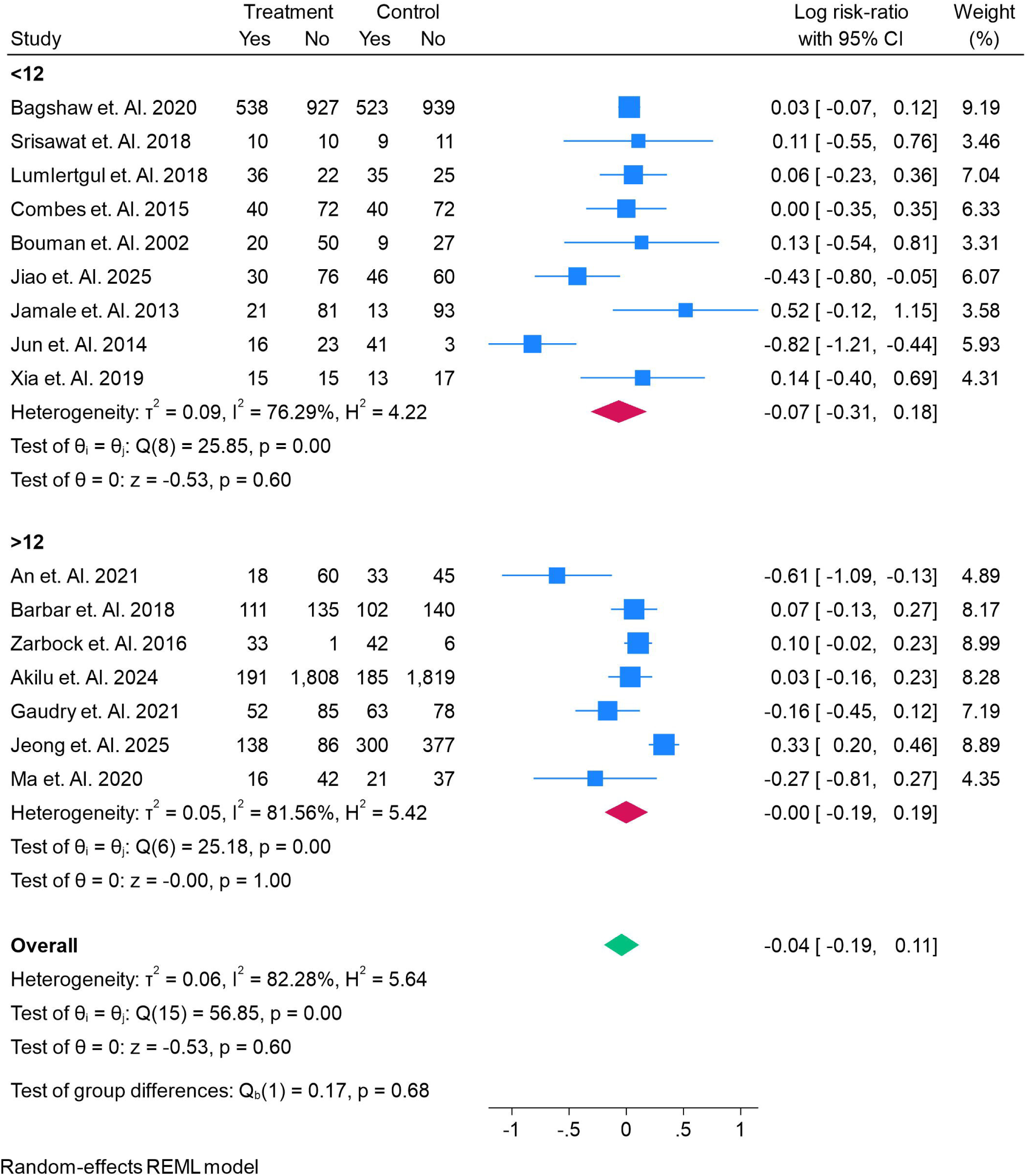
30 Days Mortality of Risk ratio in early vs delayed RRT in AKI, Sub-group analysis of SOFA.

### Subgroup Analysis of 30-Day Mortality by Age

Age-stratified analysis demonstrated no significant difference in 30-day mortality between early and delayed RRT initiation. Among patients younger than 65 years, four RCTs—including Bagshaw et al. (2020), Combes et al. (2015), Jiao et al. (2025), and Jamale et al. (2013)—reported a pooled risk ratio of 0.98 (95% CI: 0.73–1.25; p = 0.87), with moderate heterogeneity (I² = 67%). While Jamale et al. suggested a higher risk of mortality with early initiation, Jiao et al. showed a potential protective effect of delayed initiation, underscoring inconsistent results across smaller cohorts.

In patients aged ≥65 years, 12 trials—including IDEAL-ICU (Barbar et al. 2018), AKIKI (Gaudry et al. 2016), and STARRT-AKI (Bagshaw et al. 2020 subset)—showed no mortality benefit of early initiation (RR = 0.95, 95% CI: 0.76–1.14; p = 0.58), though heterogeneity remained high (I² = 83%). Some smaller studies, such as An et al. (2021) and Jeong et al. (2025), favored early initiation, whereas others including Jun et al. (2014) suggested delayed therapy may be advantageous.

The overall pooled effect across both age groups confirmed the absence of mortality benefit with early initiation (RR = 0.96, 95% CI: 0.81–1.11; p = 0.60). Furthermore, the test for subgroup differences was not significant (p = 0.86), indicating that age did not modify the association between RRT timing and short-term survival. Figure 3.

**Figure 3.**
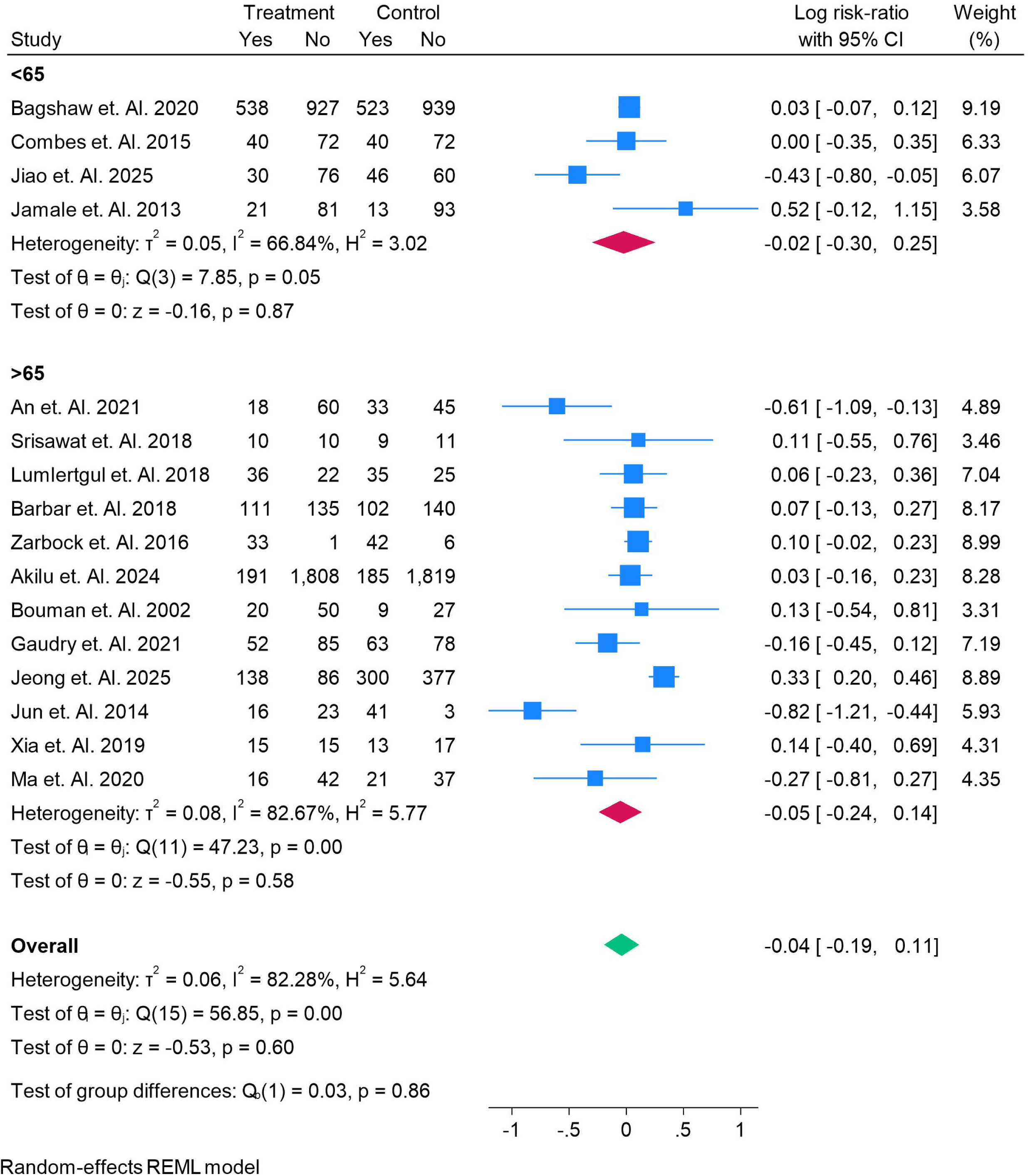
30 Days Mortality of Risk ratio in early vs delayed RRT in AKI, Sub-group analysis of Age.

### Subgroup Analysis of 90-Day Mortality by Age

Analysis of 90-day mortality stratified by age showed no significant survival advantage for early RRT initiation compared with delayed initiation. Among patients younger than 65 years, five RCTs—including Bagshaw et al. (2020), Combes et al. (2015), and Jiao et al. (2025)—reported a pooled risk ratio of 0.91 (95% CI: 0.63–1.19; p = 0.52), with moderate heterogeneity (I² = 71%). While Combes et al. suggested a modest trend favoring early initiation, Jiao et al. indicated a potential benefit for delayed initiation, reflecting inconsistent results.

In the ≥65-year subgroup, five RCTs—including IDEAL-ICU (Barbar et al. 2018), AKIKI-2 (Gaudry et al. 2021), and Jeong et al. (2025)—demonstrated no significant difference between early and delayed initiation (RR = 0.89, 95% CI: 0.72–1.07; p = 0.23), with substantial heterogeneity (I² = 75%). Of note, smaller studies such as An et al. (2021) reported lower mortality with early initiation, whereas Jeong et al. (2025) and Ma et al. (2020) showed neutral effects.

The overall pooled estimate across both age groups confirmed that early initiation did not reduce 90-day mortality (RR = 0.90, 95% CI: 0.76–1.04; p = 0.17). Moreover, the test for subgroup differences was not significant (p = 0.92), suggesting that age does not modify the impact of RRT timing on long-term outcomes. Figure 4.

**Figure 4.**
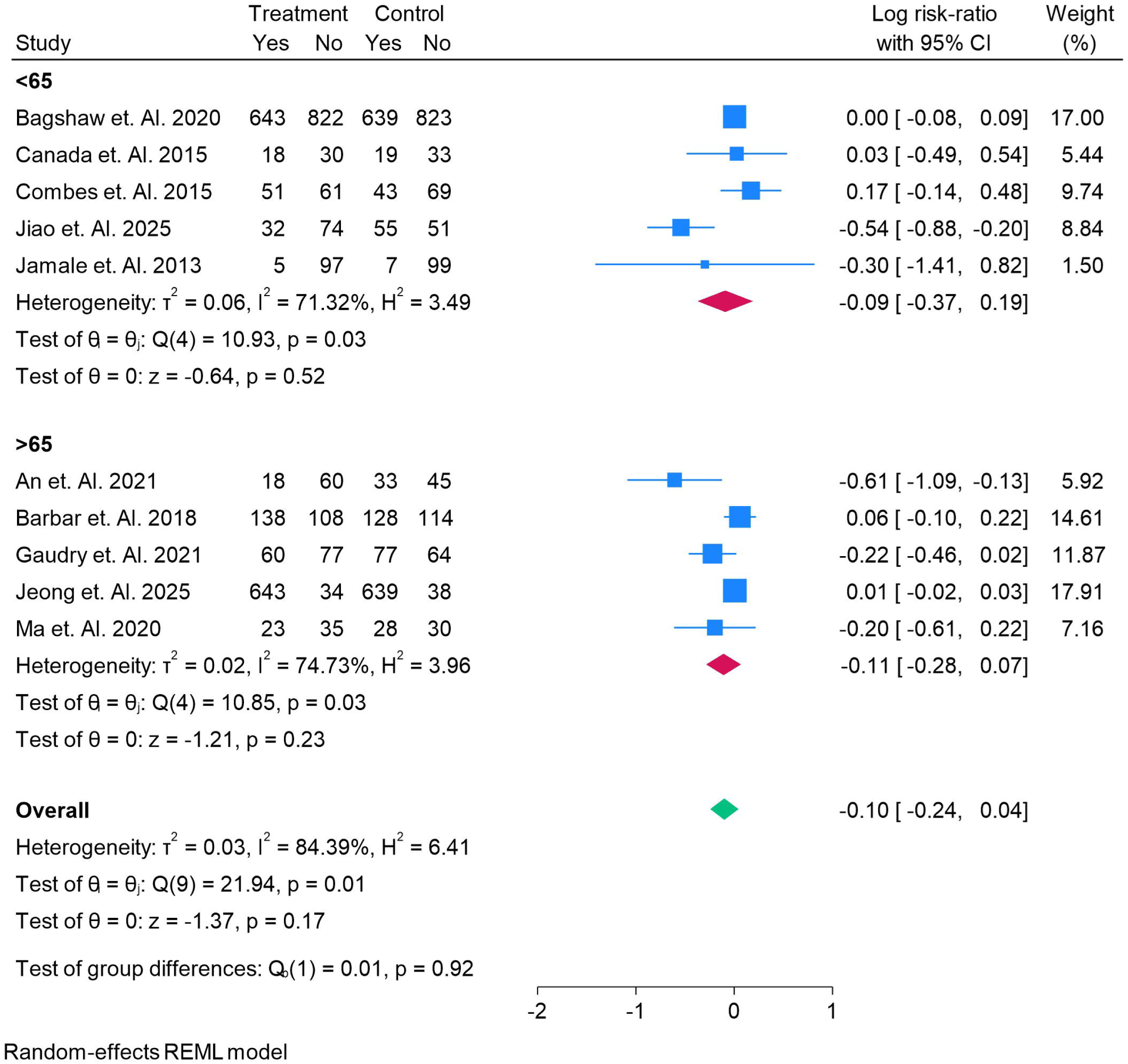
90 Days Mortality of Risk ratio in early vs delayed RRT in AKI, Sub-group analysis of AGE.

### Subgroup Analysis of 90-Day Mortality by SOFA Score

Subgroup analysis based on baseline SOFA scores showed no significant differences in 90-day mortality between early and delayed RRT initiation. In patients with SOFA <12, four RCTs—including Bagshaw et al. (2020), Combes et al. (2015), Jiao et al. (2025), and Jamale et al. (2013)—reported a pooled risk ratio of 0.88 (95% CI: 0.53–1.23; p = 0.50), with substantial heterogeneity (I² = 80%). While Jiao et al. favored delayed initiation, Jamale et al. suggested worse outcomes with early initiation, reflecting discordant findings across small cohorts.

In the SOFA >12 subgroup, six RCTs—including IDEAL-ICU (Barbar et al. 2018), AKIKI-2 (Gaudry et al. 2021), and Jeong et al. (2025)—yielded a pooled risk ratio of 0.91 (95% CI: 0.70–1.12; p = 0.26), with moderate heterogeneity (I² = 64%). Notably, An et al. (2021) suggested a survival advantage with early initiation, whereas Jeong et al. and Ma et al. (2020) demonstrated neutral or slightly worse outcomes.

The overall pooled analysis across both strata confirmed no significant mortality difference between early and delayed strategies at 90 days (RR = 0.90, 95% CI: 0.76–1.04; p = 0.17). The test for subgroup differences was not significant (p = 0.86), suggesting that baseline illness severity as measured by SOFA did not modify the association between RRT timing and long-term survival. Figure 5.

**Figure 5.**
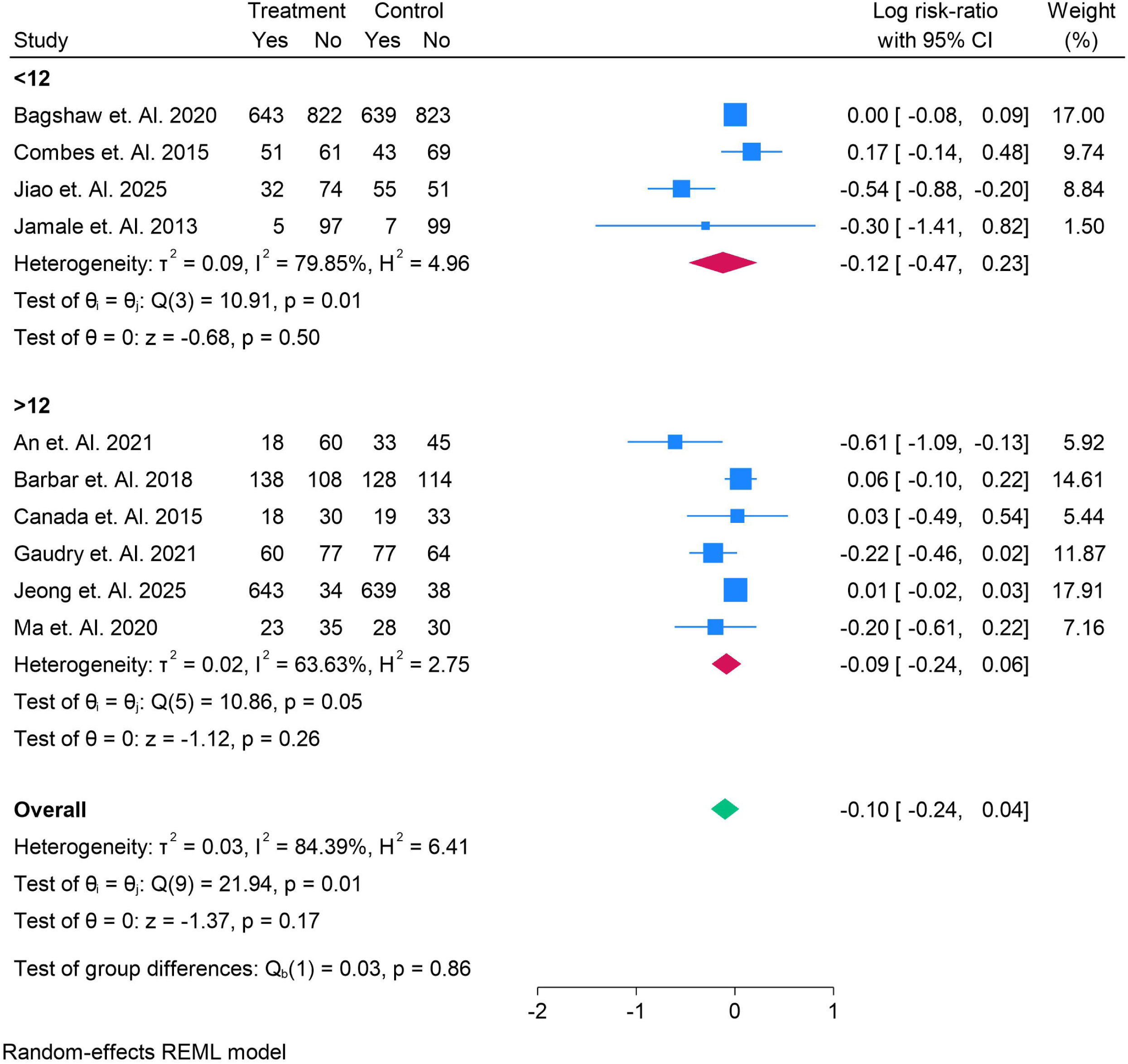
90 Days Mortality of Risk ratio in early vs delayed RRT in AKI, Sub-group analysis of SOFA.

### Subgroup Analysis of ICU Mortality by SOFA Score

When stratified by baseline SOFA scores, ICU mortality did not differ significantly between early and delayed RRT initiation. In the SOFA <12 subgroup, five RCTs—including Bagshaw et al. (2020), Combes et al. (2015), Bouman et al. (2002), Jiao et al. (2025), and Jamale et al. (2013)—yielded a pooled risk ratio of 1.06 (95% CI: 0.67–1.46; p = 0.75), with high heterogeneity (I² = 84%). While Bouman et al. suggested higher mortality with early initiation, Jiao et al. indicated better outcomes with delayed therapy, highlighting inconsistency across smaller single-center trials.

In the SOFA >12 subgroup, four RCTs—including IDEAL-ICU (Barbar et al. 2018), AKIKI-2 (Gaudry et al. 2021), and Jeong et al. (2025)—showed no survival benefit of early initiation (RR = 1.03, 95% CI: 0.88–1.18; p = 0.72), with moderate heterogeneity (I² = 56%). Some trials, such as Barbar et al., suggested worse outcomes with early initiation, whereas Gaudry et al. and Jeong et al. found neutral effects.

The overall pooled analysis confirmed that early initiation did not reduce ICU mortality compared with delayed strategies (RR = 1.02, 95% CI: 0.88–1.16; p = 0.75), with no evidence of effect modification by SOFA score (test for subgroup differences, p = 0.86). Figure 6.

**Figure 6.**
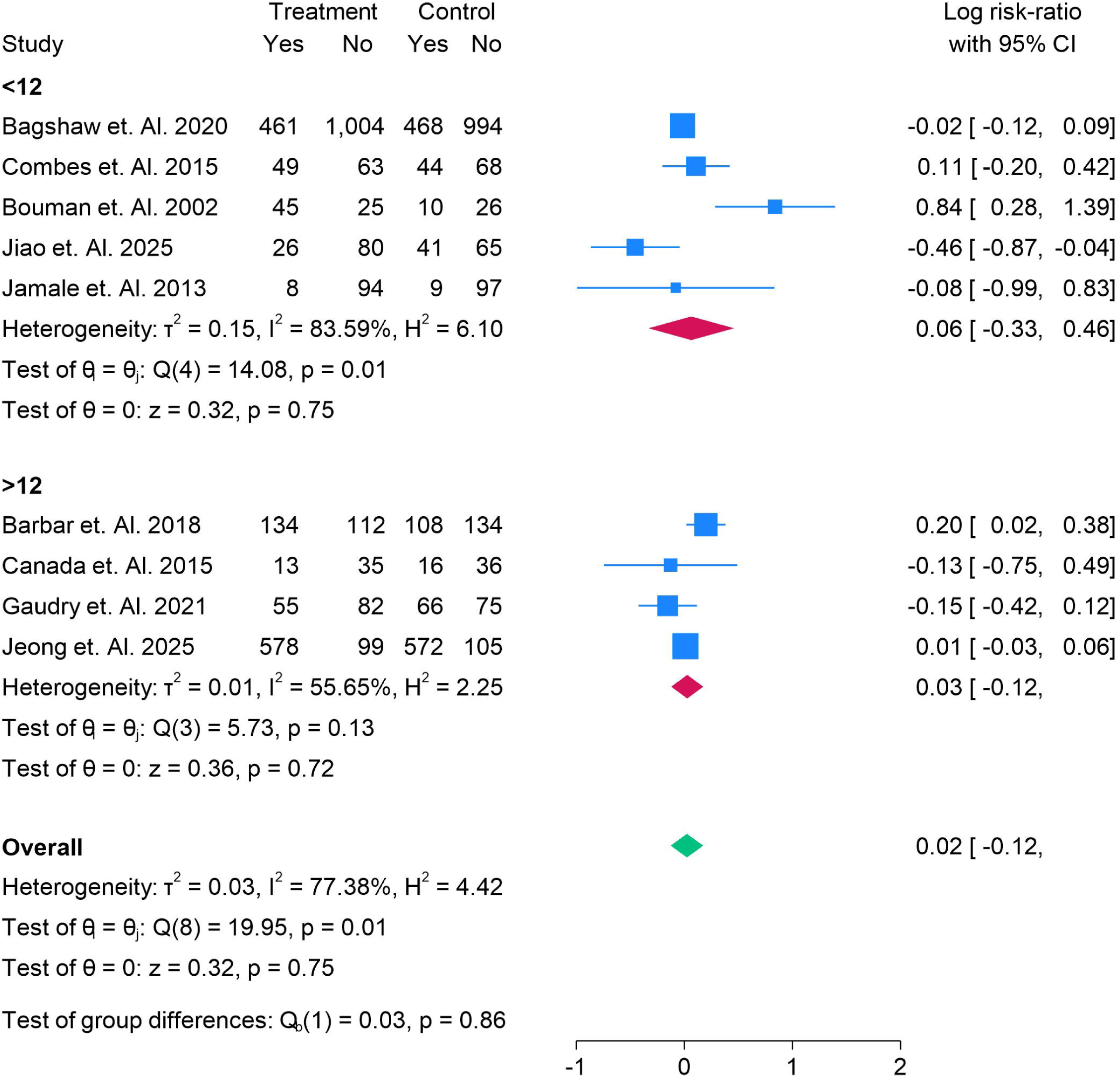
ICU Mortality of Risk ratio in early vs delayed RRT in AKI, Sub-group analysis of SOFA.

### Subgroup Analysis of ICU Mortality by Age

Age-stratified analysis of ICU mortality showed no significant benefit of early RRT initiation compared with delayed initiation. In the <65-year subgroup, five RCTs—including Bagshaw et al. (2020), Combes et al. (2015), Jiao et al. (2025), and Jamale et al. (2013)—demonstrated a pooled risk ratio of 0.94 (95% CI: 0.76–1.12; p = 0.48), with low heterogeneity (I² = 29%). While Jiao et al. suggested a survival advantage with delayed initiation, most trials reported neutral effects.

In patients aged ≥65 years, four RCTs—including IDEAL-ICU (Barbar et al. 2018), AKIKI-2 (Gaudry et al. 2021), Bouman et al. (2002), and Jeong et al. (2025)—yielded a pooled risk ratio of 1.16 (95% CI: 0.83–1.48; p = 0.35), with substantial heterogeneity (I² = 91%). Bouman et al. reported higher mortality with early initiation, whereas Gaudry et al. and Jeong et al. found no meaningful difference.

The overall pooled analysis confirmed no significant effect of early RRT on ICU mortality (RR = 1.02, 95% CI: 0.88–1.16; p = 0.75). Importantly, the test for subgroup differences was not significant (p = 0.25), suggesting that age did not modify the impact of RRT timing on ICU mortality. Figure 7.

**Figure 7.**
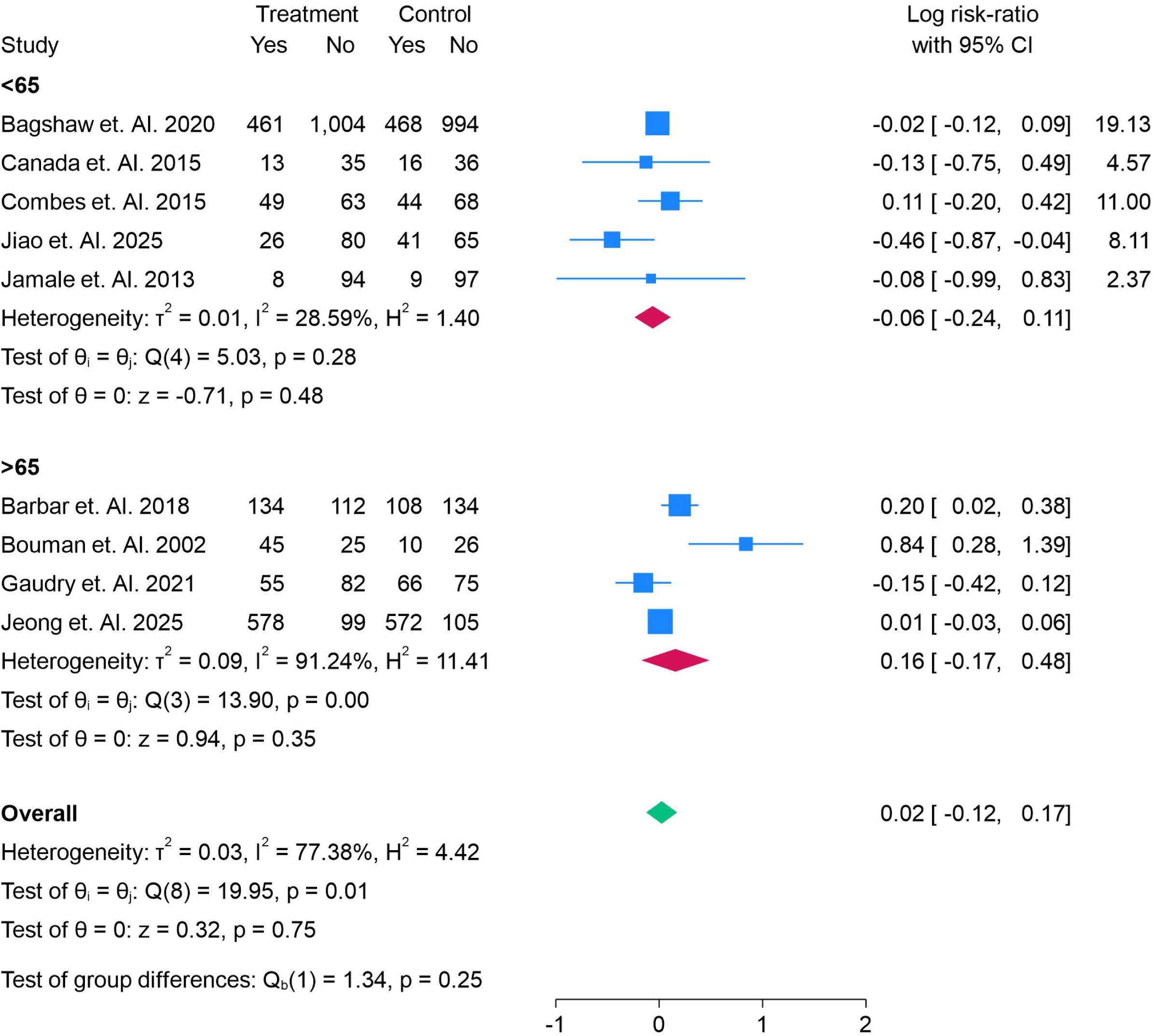
ICU Mortality of Risk ratio in early vs delayed RRT in AKI, Sub-group analysis of Age.

### Subgroup Analysis of Hospital Mortality by Age

Hospital mortality did not significantly differ between early and delayed RRT initiation when stratified by age. In the <65-year subgroup, six RCTs—including Bagshaw et al. (2020), Combes et al. (2015), Jiao et al. (2025), and Jamale et al. (2013)—demonstrated a pooled risk ratio of 0.94 (95% CI: 0.69–1.19; p = 0.64), with moderate heterogeneity (I² = 62%). While Jiao et al. reported better survival with delayed initiation, Jamale et al. indicated higher mortality with early initiation, underscoring inconsistency across smaller single-center trials.

In the ≥65-year subgroup, four RCTs—including IDEAL-ICU (Barbar et al. 2018), AKIKI-2 (Gaudry et al. 2021), Bouman et al. (2002), and Jeong et al. (2025)—showed no mortality benefit of early initiation (RR = 1.15, 95% CI: 0.56–1.46; p = 0.46), with very high heterogeneity (I² = 95%). Bouman et al. suggested worse outcomes with early initiation, whereas larger studies such as Gaudry et al. and Jeong et al. found neutral effects.

The overall pooled analysis across both age groups confirmed no significant difference in hospital mortality between early and delayed initiation (RR = 1.03, 95% CI: 0.81–1.25; p = 0.80). The test for subgroup differences was not significant (p = 0.39), indicating that age did not modify the effect of RRT timing on hospital survival. Figure 8.

**Figure 8.**
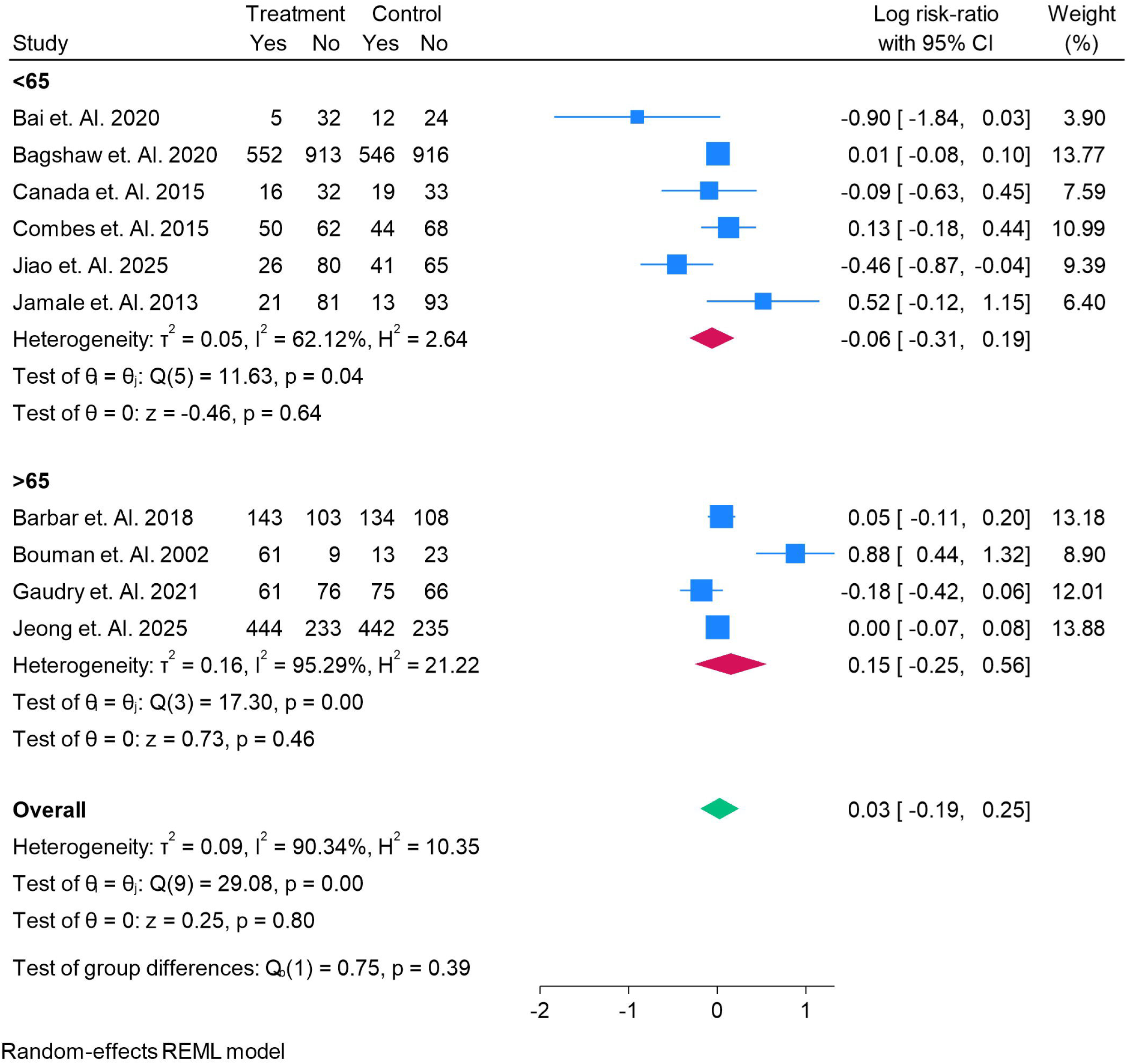
Hospital Mortality of Risk ratio in early vs delayed RRT in AKI, Sub-group analysis of Age.

### Subgroup Analysis of Hospital Mortality by SOFA Score

Hospital mortality did not significantly differ between early and delayed RRT initiation when stratified by SOFA score. In the SOFA <12 subgroup, five RCTs—including Bagshaw et al. (2020), Combes et al. (2015), Bouman et al. (2002), Jiao et al. (2025), and Jamale et al. (2013)—reported a pooled risk ratio of 1.19 (95% CI: 0.76–1.62; p = 0.39), with substantial heterogeneity (I² = 89%). Results were inconsistent: Jiao et al. suggested better survival with delayed initiation, whereas Bouman et al. indicated worse outcomes with early initiation.

In the SOFA >12 subgroup, five RCTs—including Bai et al. (2020), IDEAL-ICU (Barbar et al. 2018), AKIKI-2 (Gaudry et al. 2021), and Jeong et al. (2025)—showed no significant difference between strategies (RR = 0.99, 95% CI: 0.72–1.27; p = 0.84), with no heterogeneity (I² = 0%). Most trials in this subgroup reported neutral effects, with neither early nor delayed initiation demonstrating a consistent advantage.

The overall pooled analysis across both strata confirmed the absence of a mortality benefit with early initiation (RR = 1.03, 95% CI: 0.81–1.25; p = 0.80). The test for subgroup differences was not significant (p = 0.38), suggesting that illness severity as measured by SOFA did not influence the impact of RRT timing on hospital survival. Figure 9.

**Figure 9.**
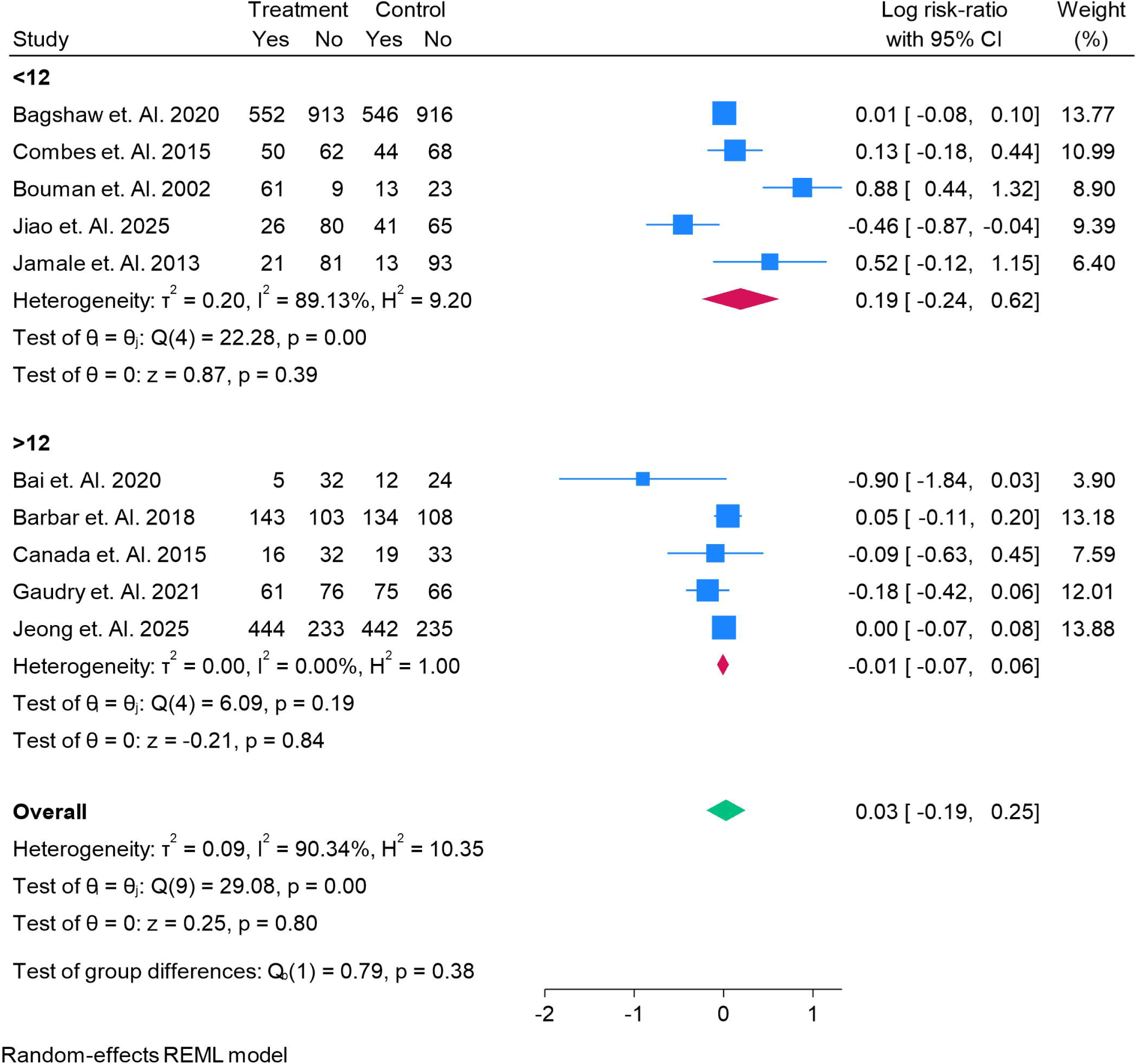
Hospital Mortality of Risk ratio in early vs delayed RRT in AKI, Sub-group analysis of SOFA.

### Subgroup Analysis of Hypotension by SOFA Score

Subgroup analysis demonstrated no significant difference in the risk of hypotension between early and delayed RRT initiation. In the SOFA <12 subgroup, five RCTs—including Bagshaw et al. (2020), Combes et al. (2015), Bouman et al. (2002), Jiao et al. (2025), and Jamale et al. (2013)—reported a pooled risk ratio of 1.19 (95% CI: 0.81–1.62; p = 0.39), with substantial heterogeneity (I² = 89%). While Jiao et al. suggested fewer hypotensive events with delayed initiation, Bouman et al. indicated more complications with early initiation, reflecting variability in smaller single-center trials.

In the SOFA >12 subgroup, five RCTs—including Bai et al. (2020), IDEAL-ICU (Barbar et al. 2018), Canada et al. (2015), AKIKI-2 (Gaudry et al. 2021), and Jeong et al. (2025)—showed no significant difference (RR = 0.99, 95% CI: 0.72–1.27; p = 0.84), with no observed heterogeneity (I² = 0%). These larger multicenter studies consistently demonstrated neutral effects.

The overall pooled analysis confirmed no association between RRT timing and the risk of hypotension (RR = 1.03, 95% CI: 0.81–1.25; p = 0.80). The test for subgroup differences was not significant (p = 0.38), indicating that illness severity, as assessed by SOFA score, did not modify the relationship between timing of RRT and hypotension risk. Figure 10.

**Figure 10.**
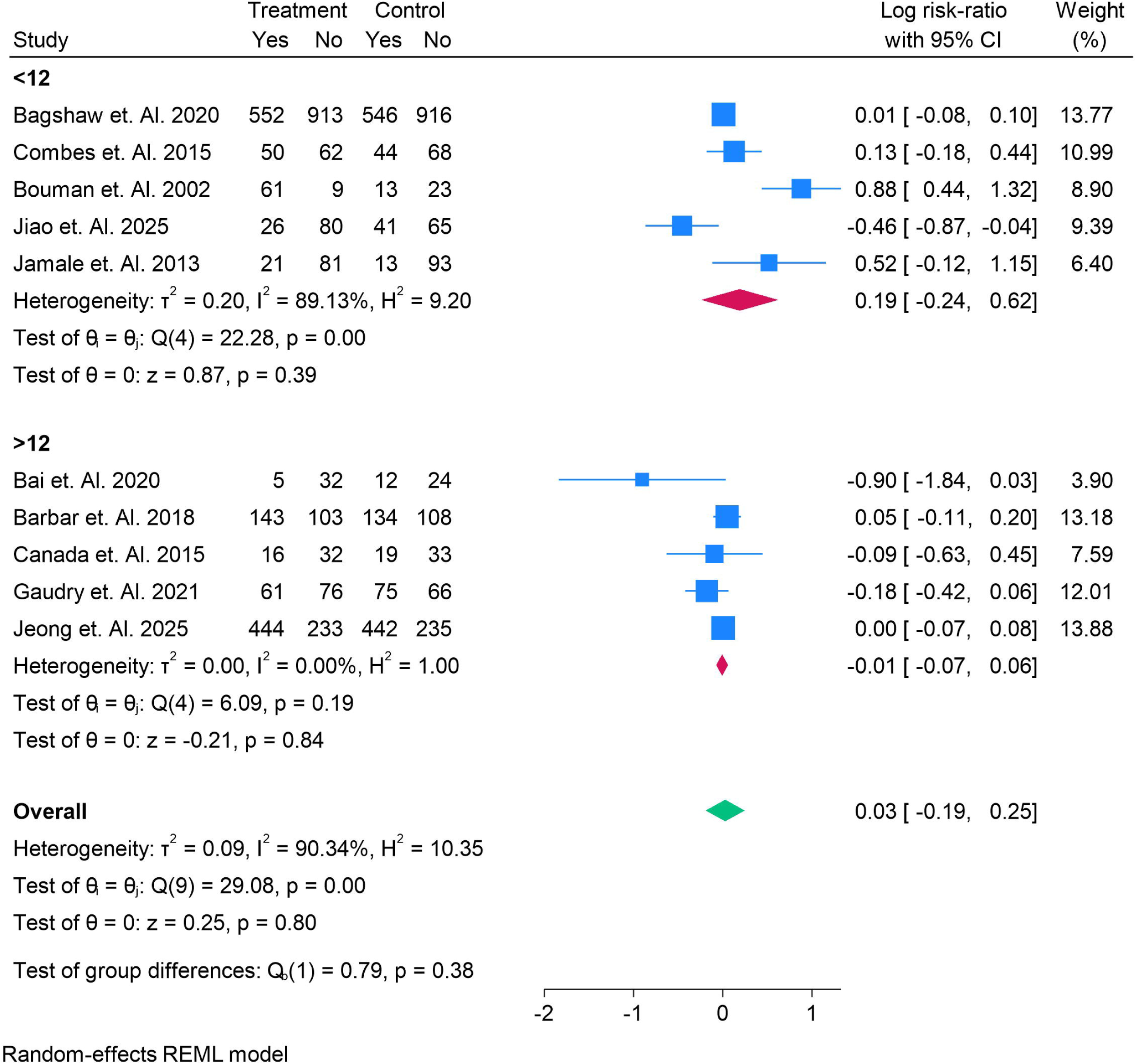
Hypotension in Risk ratio in early vs delayed RRT in AKI, Sub-group analysis of SOFA.

### Subgroup Analysis of Hypotension by Age

Age-based subgroup analysis showed no significant difference in the risk of hypotension between early and delayed RRT initiation. In the <65-year subgroup, six RCTs—including Bagshaw et al. (2020), Combes et al. (2015), Jiao et al. (2025), and Jamale et al. (2013)—reported a pooled risk ratio of 0.94 (95% CI: 0.69–1.19; p = 0.64), with moderate heterogeneity (I² = 62%). Results varied, with Jiao et al. showing lower hypotension rates in the delayed group, whereas Jamale et al. suggested more adverse events with early initiation.

In the ≥65-year subgroup, four RCTs—including IDEAL-ICU (Barbar et al. 2018), Bouman et al. (2002), AKIKI-2 (Gaudry et al. 2021), and Jeong et al. (2025)—demonstrated no significant difference between strategies (RR = 1.15, 95% CI: 0.56–1.46; p = 0.46), with very high heterogeneity (I² = 95%). Bouman et al. suggested increased hypotension risk with early initiation, whereas larger trials such as Gaudry et al. and Jeong et al. reported neutral outcomes.

The overall pooled analysis across both subgroups confirmed no significant difference in hypotension risk between early and delayed initiation (RR = 1.03, 95% CI: 0.81–1.25; p = 0.80). The test for subgroup differences was not significant (p = 0.39), indicating that age did not modify the effect of RRT timing on hypotension risk. Figure 11.

**Figure 11.**
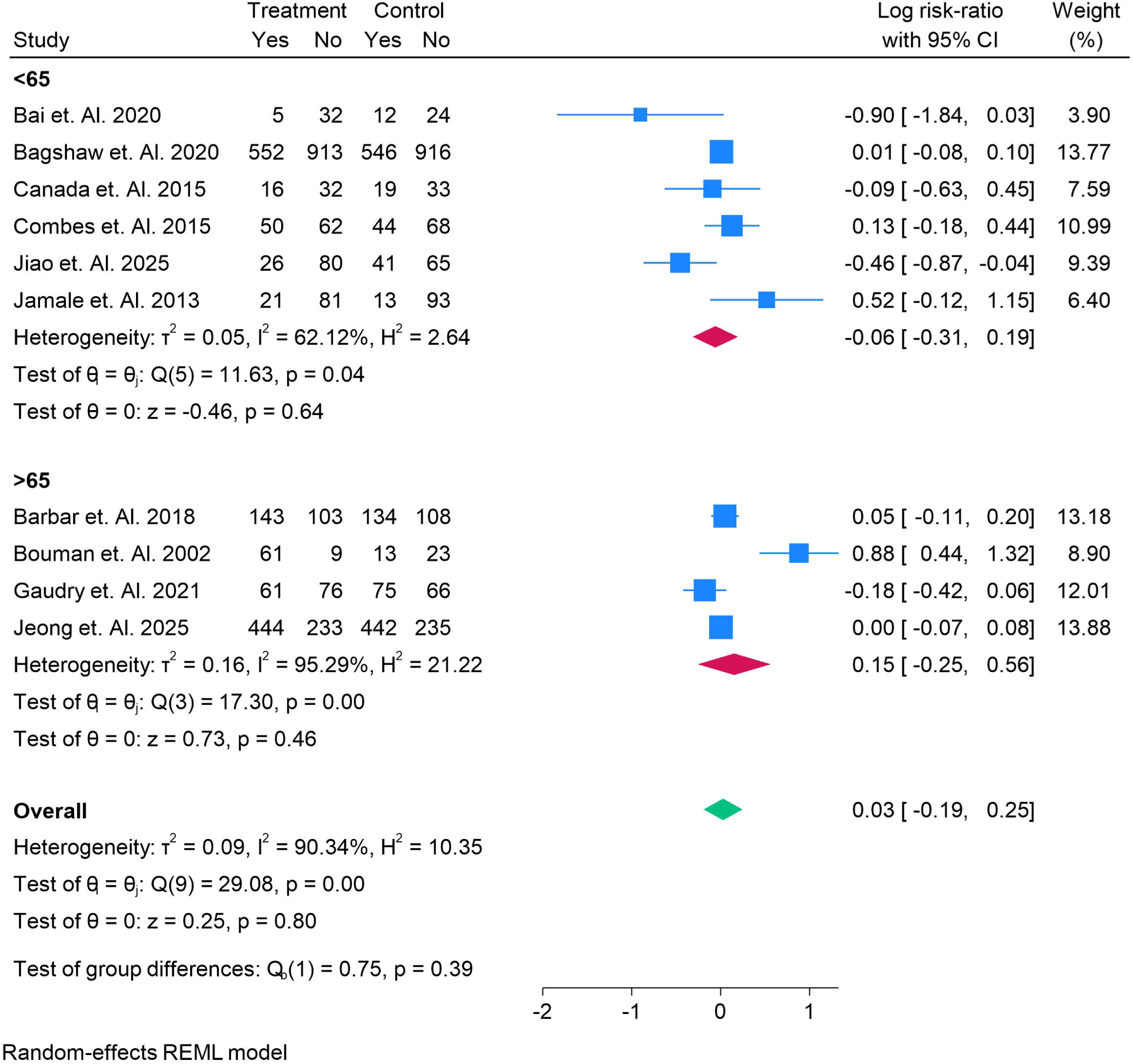
Hypotension in Risk ratio in early vs delayed RRT in AKI, Sub-group analysis of Age.

### Subgroup Analysis of Infection by Age

Infection rates were significantly higher in patients receiving early RRT initiation compared with delayed initiation. In the <65-year subgroup, four RCTs—including Bagshaw et al. (2020), Combes et al. (2015), Canada et al. (2015), and Jiao et al. (2025)—demonstrated a pooled risk ratio of 1.15 (95% CI: 0.69–1.61; p = 0.52), with considerable heterogeneity (I² = 83%). While Bagshaw et al. reported increased risk of bloodstream infections with early initiation, Jiao et al. suggested a protective effect with delayed therapy.

In the ≥65-year subgroup, two RCTs—IDEAL-ICU (Barbar et al. 2018) and Jeong et al. (2025)—reported a pooled risk ratio of 1.23 (95% CI: 0.98–1.47; p = 0.07), with moderate heterogeneity (I² = 67%). Both trials showed a tendency toward increased infections in the early group, though results did not reach statistical significance individually.

The overall pooled analysis confirmed that early initiation was associated with a higher risk of infection compared with delayed initiation (RR = 1.21, 95% CI: 1.03–1.39; p = 0.02). The test for subgroup differences was not significant (p = 0.78), suggesting that the increased infection risk with early initiation was consistent across both younger and older patients. Figure 12.

**Figure 12.**
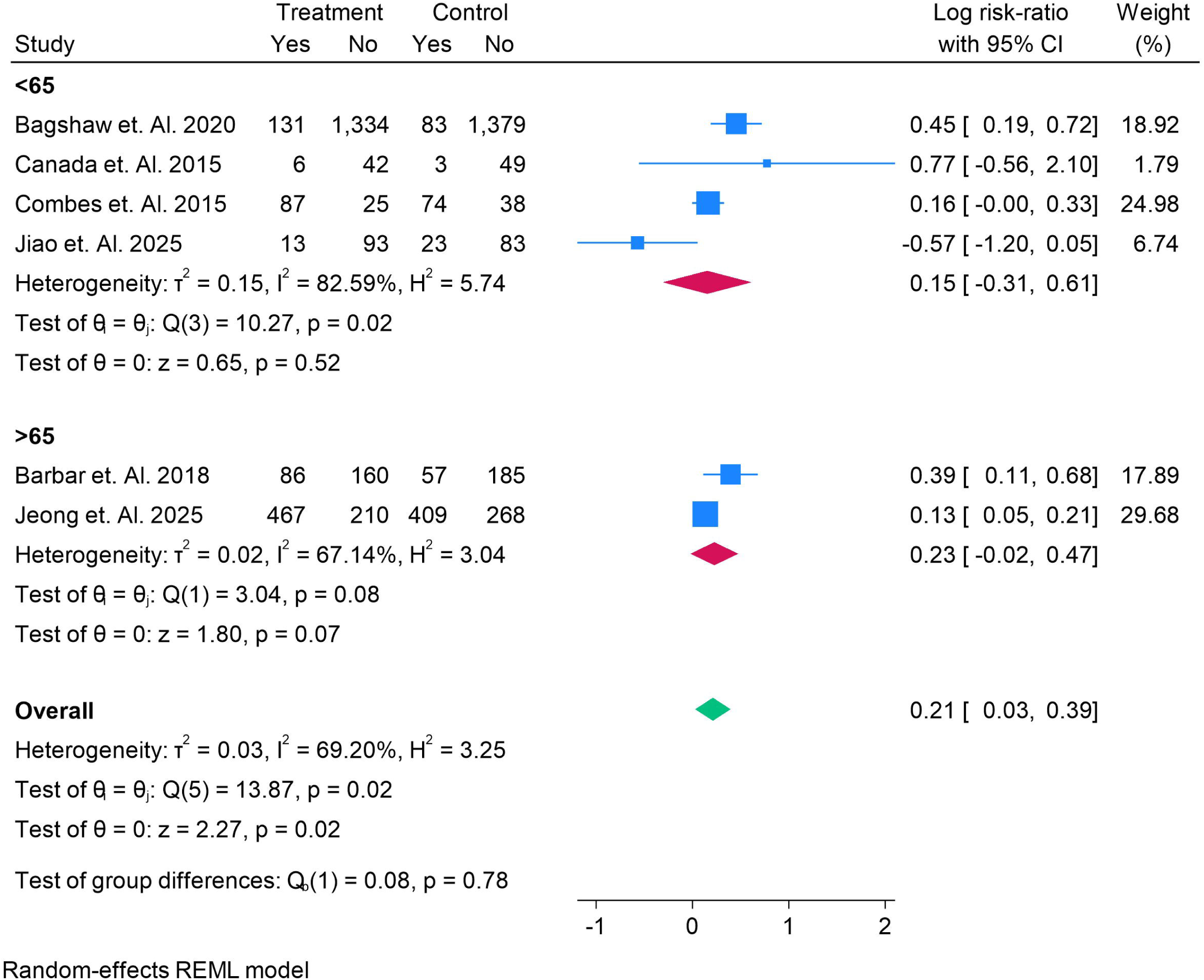
Infection in Risk ratio in early vs delayed RRT in AKI, Sub-group analysis of Age.

### Subgroup Analysis of Infection by SOFA Score

When stratified by baseline SOFA score, early initiation of RRT was associated with a higher risk of infection compared with delayed initiation. In the SOFA <12 subgroup, three RCTs—Bagshaw et al. (2020), Combes et al. (2015), and Jiao et al. (2025)—reported a pooled risk ratio of 1.08 (95% CI: 0.56–1.60; p = 0.76), with high heterogeneity (I² = 89%). While Bagshaw et al. and Combes et al. demonstrated increased infection risk in the early group, Jiao et al. showed a possible benefit with delayed initiation, reflecting variability across studies.

In the SOFA >12 subgroup, three RCTs—IDEAL-ICU (Barbar et al. 2018), Canada et al. (2015), and Jeong et al. (2025)—reported a pooled risk ratio of 1.25 (95% CI: 1.00–1.49; p = 0.05), with moderate heterogeneity (I² = 53%). Both Barbar et al. and Jeong et al. suggested a significantly higher infection risk in the early initiation group.

The overall pooled analysis confirmed that early initiation of RRT was associated with an increased risk of infection (RR = 1.21, 95% CI: 1.03–1.39; p = 0.02). The test for subgroup differences was not significant (p = 0.58), indicating that the elevated infection risk with early initiation was consistent regardless of baseline illness severity. Figure 13.

**Figure 13.**
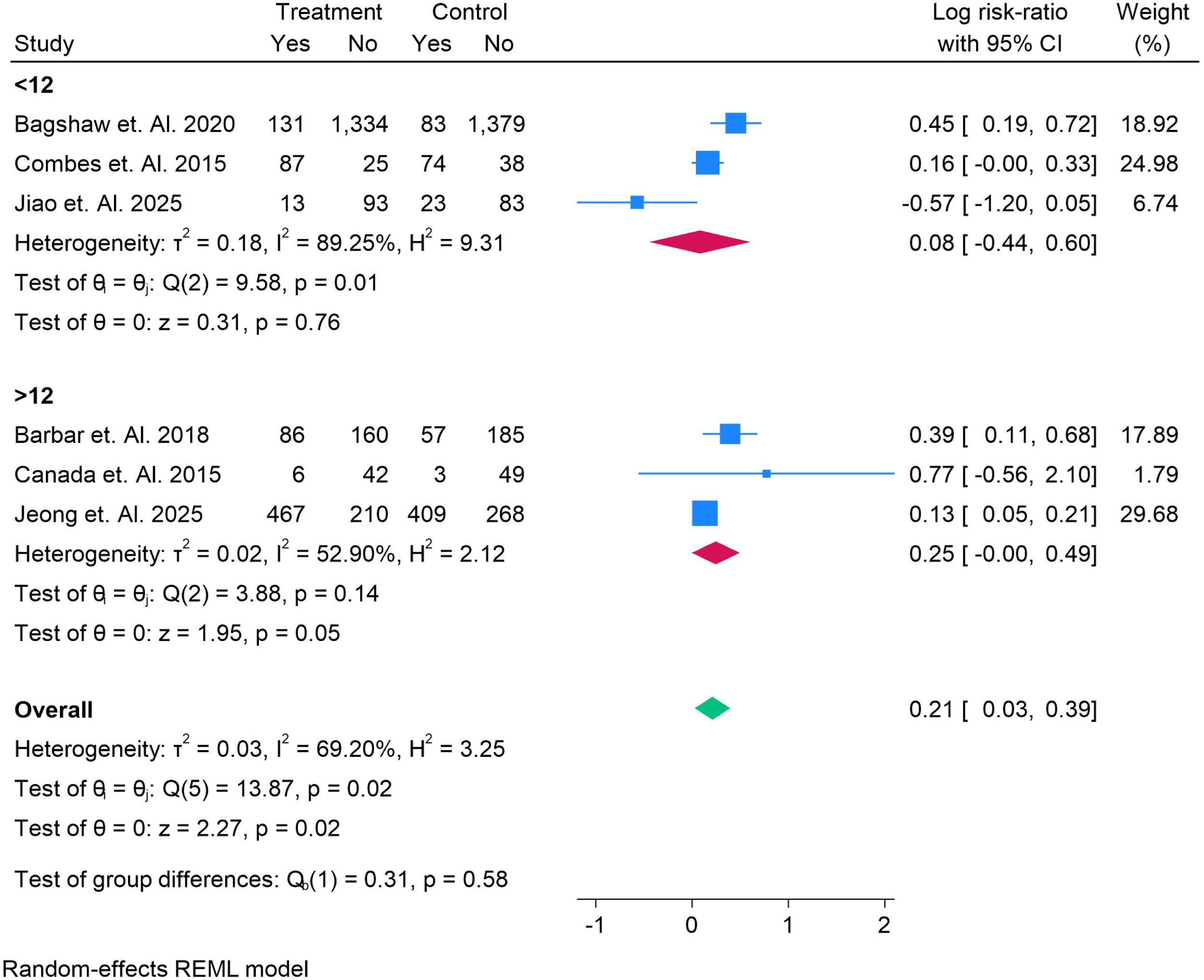
Infection in Risk ratio in early vs delayed RRT in AKI, Sub-group analysis of SOFA.

### Subgroup Analysis of Arrhythmia by SOFA Score

Arrhythmia risk did not significantly differ between early and delayed RRT initiation when stratified by baseline SOFA scores. In the SOFA <12 subgroup, two RCTs—STARRT-AKI (Bagshaw et al. 2020) and AKIKI (Gaudry et al. 2016)—reported a pooled risk ratio of 1.20 (95% CI: 0.67–1.73; p = 0.49), with moderate heterogeneity (I² = 68%). Bagshaw et al. showed a trend toward higher arrhythmia incidence in the early group, whereas Gaudry et al. reported no difference.

In the SOFA >12 subgroup, six RCTs—including IDEAL-ICU (Barbar et al. 2018), Zarbock et al. (2016), AKIKI-2 (Gaudry et al. 2021), Jeong et al. (2025), and smaller single-center studies (Canada et al. 2015; Iyem et al. 2009)—demonstrated a pooled risk ratio of 1.18 (95% CI: 0.72–1.64; p = 0.94), with substantial heterogeneity (I² = 74%). While Barbar et al. and Jeong et al. suggested slightly higher arrhythmia rates in early initiation, Gaudry et al. (2021) indicated lower risk, showing inconsistent results across trials.

The overall pooled analysis confirmed no significant effect of early initiation on arrhythmia risk (RR = 1.18, 95% CI: 0.79–1.57; p = 0.54), with no evidence of subgroup modification by SOFA score (p = 0.64). Figure 14.

**Figure 14.**
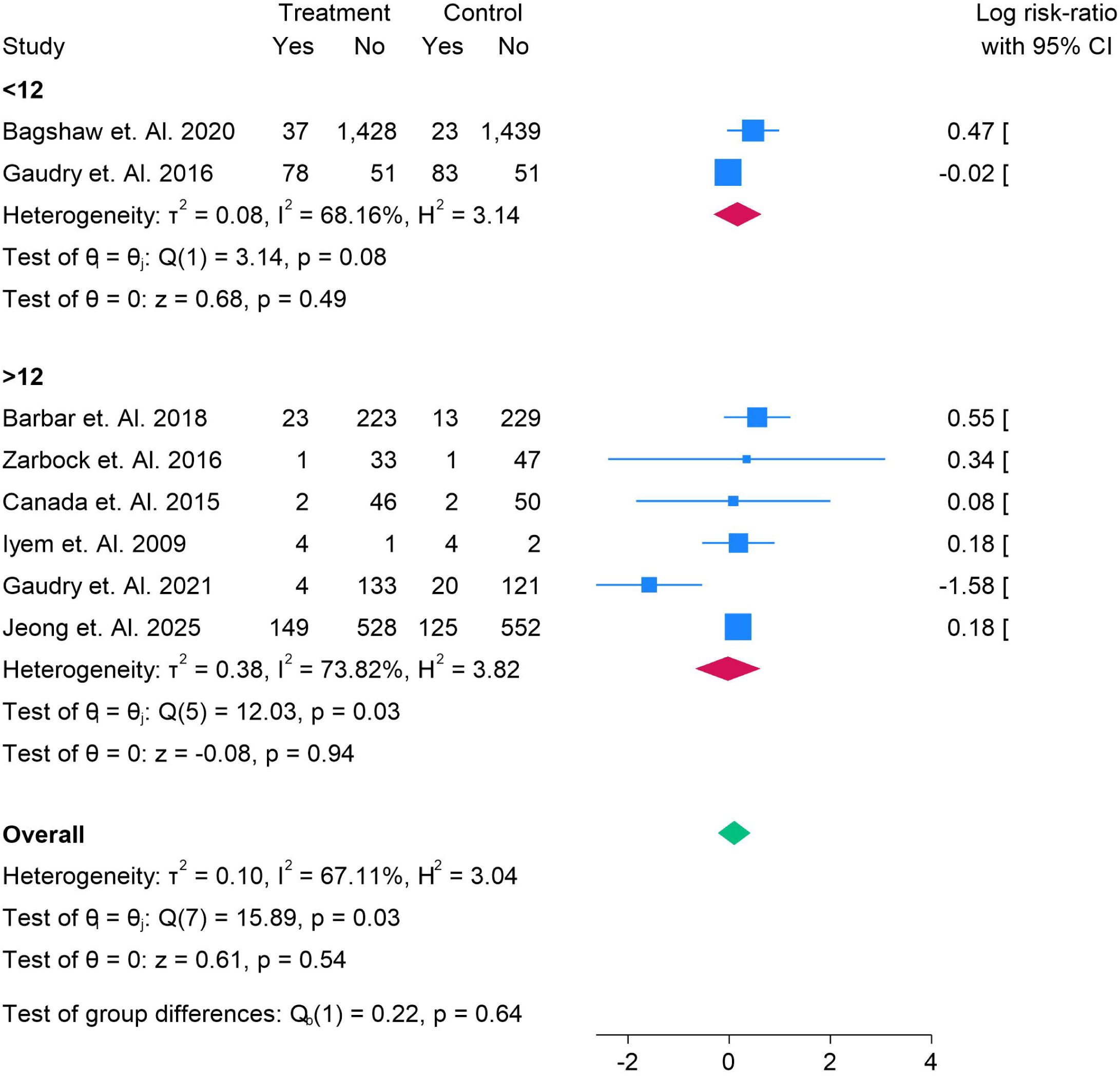
Arrythmia in Risk ratio in early vs delayed RRT in AKI, Sub-group analysis of SOFA.

### Subgroup Analysis of Arrhythmia by Age

Arrhythmia events did not significantly differ between early and delayed RRT initiation across age subgroups. In patients <65 years, four RCTs—including Bagshaw et al. (2020), Gaudry et al. (2016), Iyem et al. (2009), and Canada et al. (2015)—reported a pooled risk ratio of 1.13 (95% CI: 0.82–1.45; p = 0.41), with low heterogeneity (I² = 32%). Bagshaw et al. suggested a trend toward increased arrhythmias with early initiation, while Gaudry et al. found no difference, and smaller studies were inconclusive due to limited sample sizes.

In the ≥65-year subgroup, four RCTs—including IDEAL-ICU (Barbar et al. 2018), AKIKI-2 (Gaudry et al. 2021), Zarbock et al. (2016), and Jeong et al. (2025)—demonstrated no significant effect of timing (RR = 0.86, 95% CI: 0.57–1.15; p = 0.79), with high heterogeneity (I² = 86%). Gaudry et al. reported fewer arrhythmias with early initiation, whereas Barbar et al. and Jeong et al. showed a modest increase, indicating inconsistent findings across larger trials.

The overall pooled analysis confirmed that early initiation was not associated with an increased risk of arrhythmias compared with delayed initiation (RR = 1.10, 95% CI: 0.78–1.42; p = 0.54). The test for subgroup differences was not significant (p = 0.62), suggesting that age did not modify the effect of RRT timing on arrhythmia risk. Figure 15.

**Figure 15.**
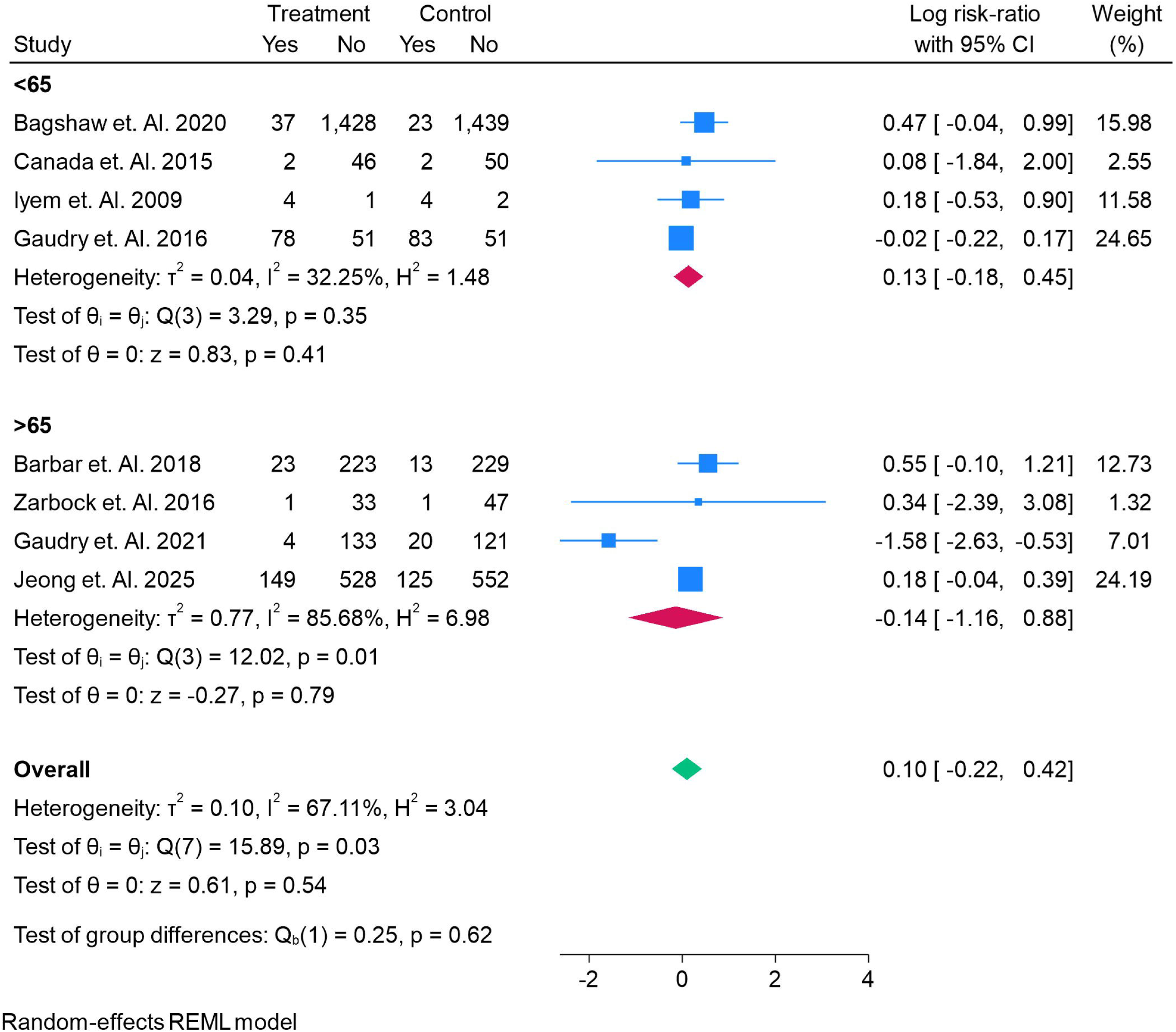
Arrythmia in Risk ratio in early vs delayed RRT in AKI, Sub-group analysis of AGE.

### Subgroup Analysis of Bleeding Events by SOFA Score

The risk of bleeding events was not significantly different between early and delayed RRT initiation in either SOFA subgroup. In the SOFA <12 subgroup, five RCTs—including Bagshaw et al. (2020), Combes et al. (2015), Gaudry et al. (2016), Bouman et al. (2002), and Jamale et al. (2013)—showed a pooled risk ratio of 0.85 (95% CI: 0.51–1.19; p = 0.39), with low heterogeneity (I² = 33%). While Gaudry et al. and Bouman et al. reported fewer bleeding events with early initiation, Bagshaw et al. noted slightly more events, though findings were inconsistent and individually underpowered.

In the SOFA >12 subgroup, four RCTs—including IDEAL-ICU (Barbar et al. 2018), Canada et al. (2015), AKIKI-2 (Gaudry et al. 2021), and Jeong et al. (2025)—demonstrated no difference between strategies (RR = 1.02, 95% CI: 0.81–1.23; p = 0.84), with no heterogeneity (I² = 0%). Jeong et al., the largest contributor in this subgroup, reported nearly identical bleeding rates across early and delayed groups.

The overall pooled estimate confirmed that early initiation did not increase bleeding complications compared with delayed initiation (RR = 0.93, 95% CI: 0.75–1.11; p = 0.46). The test for subgroup differences was not significant (p = 0.39), indicating that baseline SOFA score did not modify bleeding risk. Figure 16.

**Figure 16.**
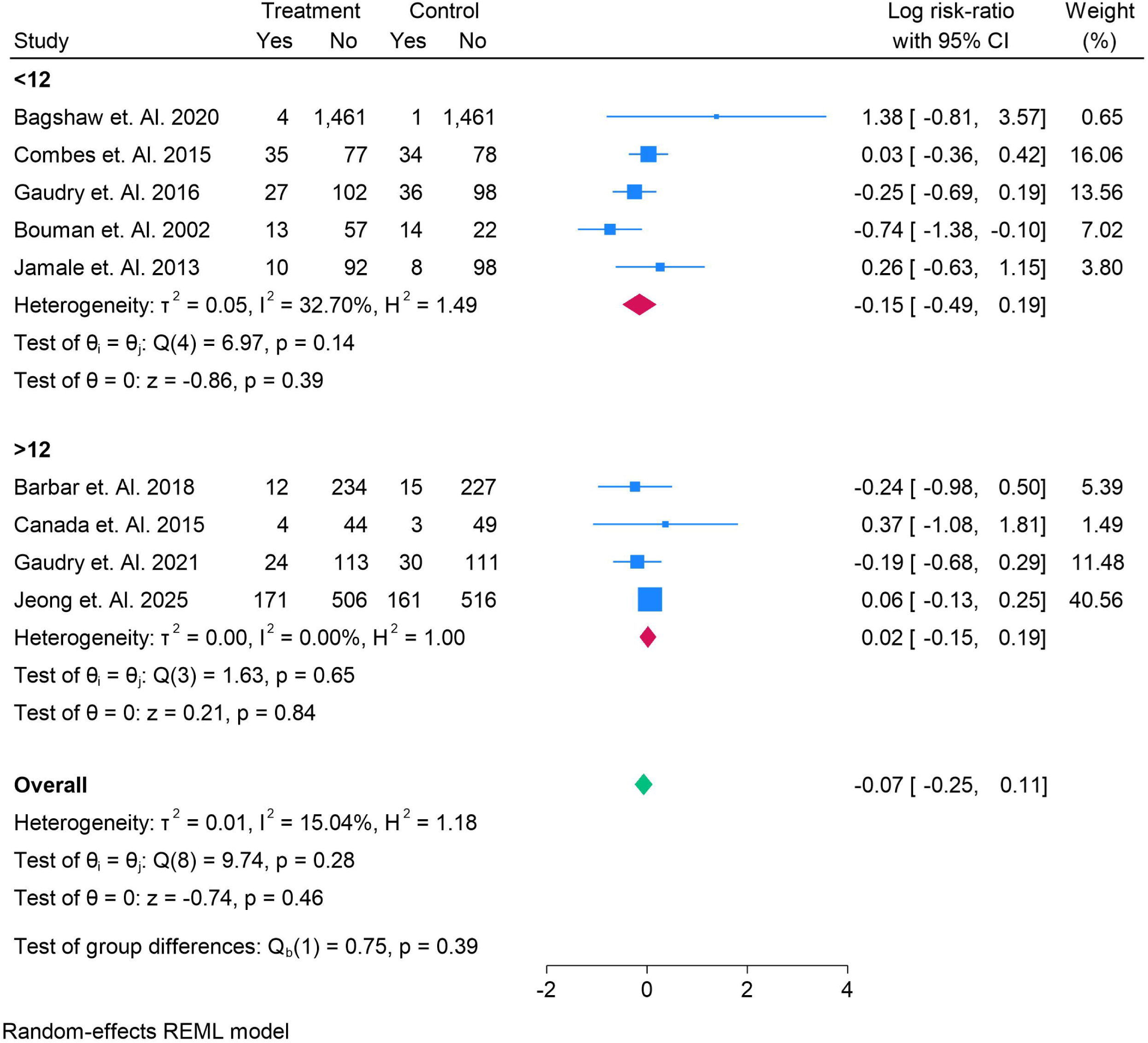
Bleeding Events in Risk ratio in early vs delayed RRT in AKI, Sub-group analysis of SOFA.

### Subgroup Analysis of Bleeding Events by Age

Bleeding complications did not significantly differ between early and delayed RRT initiation across age groups. In the <65-year subgroup, five RCTs—including Bagshaw et al. (2020), Combes et al. (2015), Gaudry et al. (2016), Jamale et al. (2013), and Canada et al. (2015)—showed a pooled risk ratio of 0.98 (95% CI: 0.71–1.25; p = 0.86), with no heterogeneity (I² = 0%). Most individual studies demonstrated neutral findings, with Gaudry et al. reporting slightly fewer bleeding events in the early group, but without statistical significance.

In the ≥65-year subgroup, four RCTs—including IDEAL-ICU (Barbar et al. 2018), AKIKI-2 (Gaudry et al. 2021), Bouman et al. (2002), and Jeong et al. (2025)—yielded a pooled risk ratio of 0.81 (95% CI: 0.47–1.15; p = 0.27), with moderate heterogeneity (I² = 53%). Bouman et al. reported fewer bleeding events in the early group, whereas Jeong et al. found nearly identical risks between strategies.

The overall pooled estimate confirmed no difference in bleeding events between early and delayed RRT initiation (RR = 0.93, 95% CI: 0.75–1.11; p = 0.46). The test for subgroup differences was not significant (p = 0.45), suggesting that age did not influence bleeding risk related to timing of RRT. Figure 17.

**Figure 17.**
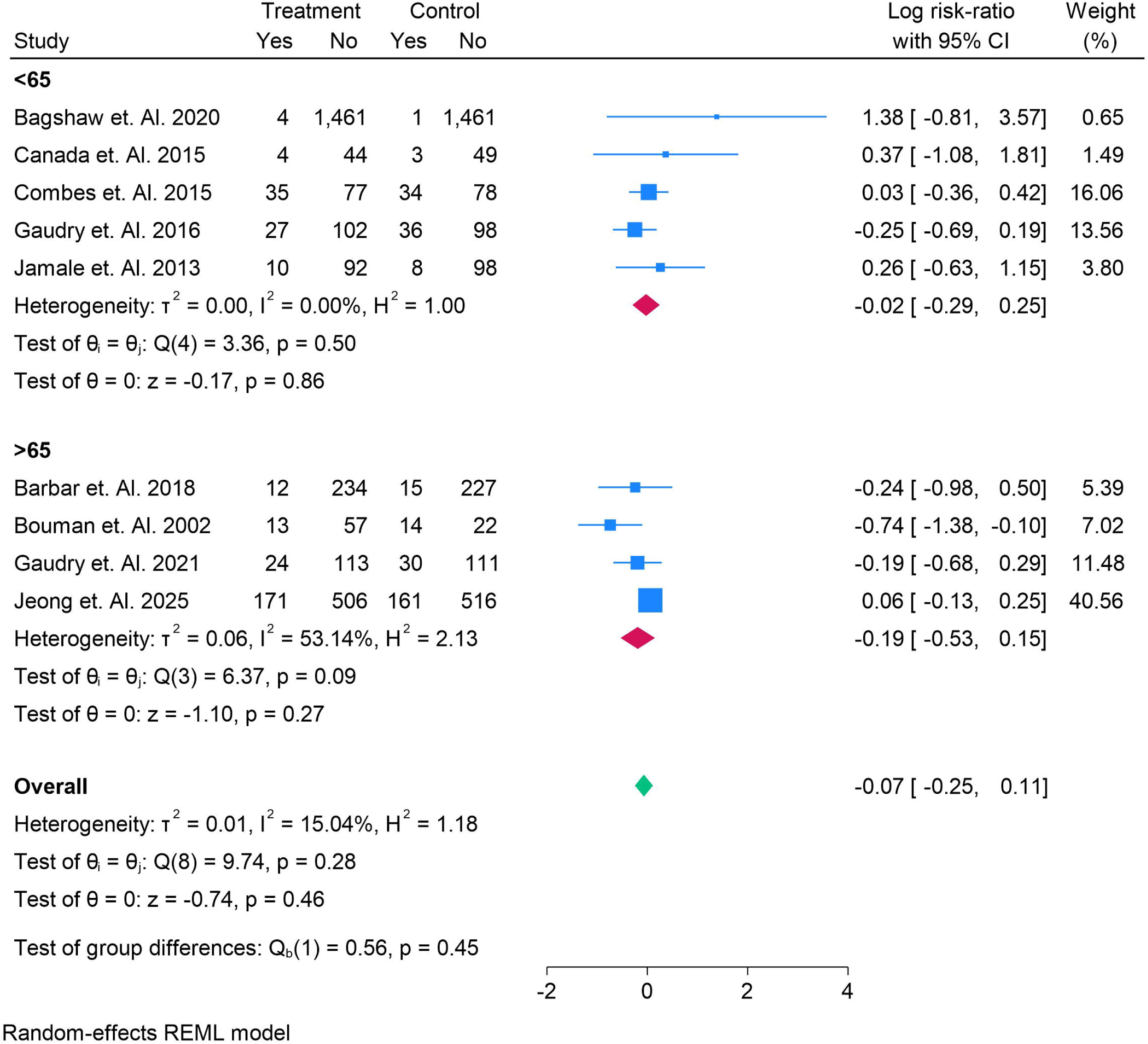
Bleeding Events in Risk ratio in early vs delayed RRT in AKI, Sub-group analysis of AGE.

## Discussion

In this updated systematic review and meta-analysis of randomized controlled trials (RCTs), we found that **early initiation of renal replacement therapy (RRT) in critically ill patients with acute kidney injury (AKI) did not significantly reduce short-term or long-term mortality compared with delayed initiation**. Across subgroup analyses stratified by age and severity of illness (SOFA score), early initiation was not associated with survival benefit at 30 days, 90 days, ICU discharge, or hospital discharge. Importantly, early initiation was linked to a **higher risk of infection**, while risks of hypotension, arrhythmia, and bleeding events did not differ between strategies. Taken together, our findings reinforce that **a “watchful waiting” strategy remains safe and avoids unnecessary RRT exposure in patients who may recover kidney function spontaneously**, without compromising mortality outcomes.

### Comparison with Previous Evidence

Our results are consistent with the largest multicenter RCT to date, the **STARRT-AKI trial**, which enrolled over 3,000 patients and demonstrated no mortality benefit with accelerated RRT initiation, but reported increased RRT dependence in survivor [31]. Similarly, the **AKIKI trial** found no difference in 60-day mortality between early and delayed initiation, with nearly half of patients in the delayed group never requiring RRT, underscoring the importance of avoiding unnecessary interventions [35]. The **IDEAL-ICU trial**, focusing on septic shock–associated AKI, was stopped early for futility, again showing no mortality advantage with early initiation [20].

In contrast, the **ELAIN trial**, a single-center study with 231 patients, suggested that early initiation may reduce 90-day mortality [21]. However, differences in patient population (post-cardiac surgery patients with lower baseline SOFA scores) and trial design (stricter timing definitions) may explain the discrepant findings. Our subgroup analyses support this interpretation: in less severely ill patients (SOFA <12), some smaller studies suggested trends favoring early initiation, whereas in more severely ill patients (SOFA ≥12), early initiation consistently failed to improve survival.

Several prior meta-analyses have addressed this question, but findings have been inconsistent due to heterogeneity in study populations, RRT modalities, and definitions of “early” versus “delayed.” A 2018 meta-analysis by Moreira et al. concluded that early initiation did not improve survival and increased the risk of complications [36]. A recent meta-analysis by Li et al. (2023), incorporating newer RCTs, similarly reported no mortality benefit but highlighted increased adverse events with early initiation [37]. Our study builds on this evidence by performing detailed subgroup analyses by SOFA and age, confirming that neither baseline illness severity nor age modifies the effect of RRT timing on mortality.

### Adverse Events and Safety Outcomes

While mortality was unaffected, our analysis found that **early initiation increased the risk of infection**, consistent with the hypothesis that unnecessary exposure to invasive vascular access and extracorporeal circuits predisposes to bloodstream infections. Both STARRT-AKI and IDEAL-ICU reported higher rates of catheter-related bloodstream infections in the early groups [31,10]. Conversely, risks of **hypotension, arrhythmia, and bleeding events** were not significantly different between strategies in our pooled analyses, suggesting that the hemodynamic and coagulation risks of RRT are similar once patients are initiated, regardless of timing.

### Clinical Implications

The timing of RRT initiation remains one of the most debated issues in critical care nephrology. Our findings support a **delayed or conservative approach**, reserving RRT for patients with urgent life-threatening complications (e.g., refractory hyperkalemia, metabolic acidosis, or fluid overload) or persistent oliguria with worsening azotemia. This strategy prevents unnecessary initiation in patients likely to recover renal function, reduces RRT exposure, and avoids infection risk, without adversely affecting mortality.

Importantly, nearly half of patients randomized to the delayed arm in trials such as AKIKI and IDEAL-ICU never required RRT [31,10]. This underscores the potential for overtreatment in early strategies and the importance of individualized clinical judgment. Furthermore, resource utilization is a major consideration: delaying RRT in appropriate patients may optimize ICU resource allocation without harming outcomes.

### Limitations

Several limitations should be acknowledged. First, although our meta-analysis included large multicenter RCTs, heterogeneity in timing definitions persists; “early” initiation ranged from within 6 hours of eligibility in ELAIN to within 12 hours in STARRT-AKI, while “delayed” initiation varied from waiting for emergent indications to 48–72 hours. Second, subgroup analyses by SOFA and age are limited by the smaller sample sizes within individual strata, which may reduce precision. Third, long-term outcomes such as quality of life, renal recovery beyond 90 days, and cost-effectiveness were not consistently reported across trials. Finally, while publication bias was not evident, the possibility of small-study effects cannot be excluded.

### Conclusions

This updated meta-analysis demonstrates that **early initiation of RRT in critically ill patients with AKI does not improve survival, regardless of age or baseline illness severity**. Early initiation, however, is associated with an increased risk of infection without reducing other adverse events. Our findings support a **delayed initiation strategy**, reserving RRT for patients with absolute indications or those failing conservative management, in alignment with the growing body of high-quality evidence. Future research should focus on identifying biomarkers or predictive tools to better stratify patients most likely to benefit from early initiation while sparing others unnecessary treatment.

## Authors’ contributions

All authors contributed equally to the manuscript and read and approved the final version of the manuscript.

## Data Availability

Supplementary file

## References

1. Mo S, Bjelland TW, Nilsen TIL, Klepstad P. Acute kidney injury in intensive care patients: Incidence, time course, and risk factors. Acta Anaesthesiol Scand. 2022 Sep;66(8):961–968. doi: 10.1111/aas.14100. Epub 2022 Jun 26. PMID: 35674748; PMCID: PMC9543500.

2. Lafrance JP, Miller DR. Acute kidney injury associates with increased long-term mortality. J Am Soc Nephrol. 2010 Feb;21(2):345–52. doi: 10.1681/ASN.2009060636. Epub 2009 Dec 17. PMID: 20019168; PMCID: PMC2834549.

3. Lee SA, Cozzi M, Bush EL, Rabb H. Distant Organ Dysfunction in Acute Kidney Injury: A Review. Am J Kidney Dis. 2018 Dec;72(6):846–856. doi: 10.1053/j.ajkd.2018.03.028. Epub 2018 Jun 14. PMID: 29866457; PMCID: PMC6252108.

4. Romagnoli S, Ricci Z. When to start a renal replacement therapy in acute kidney injury (AKI) patients: many irons in the fire. Ann Transl Med. 2016 Sep;4(18):355. doi: 10.21037/atm.2016.08.55. PMID: 27761459; PMCID: PMC5066036.

5. Ko JP, Ng LS, Goh KJ, Chai HZ, Phua GC, Tan QL. Staff perception and attitudes towards a medical rapid response team with a multi-tiered response. Singapore Med J. 2023 Aug;64(8):527–533. doi: 10.11622/smedj.2021223. PMID: 34911185; PMCID: PMC10476913.

6. Castro I, Relvas M, Gameiro J, Lopes JA, Monteiro-Soares M, Coentrão L. The impact of early versus late initiation of renal replacement therapy in critically ill patients with acute kidney injury on mortality and clinical outcomes: a meta-analysis. Clin Kidney J. 2022 May 12;15(10):1932–1945. doi: 10.1093/ckj/sfac139. PMID: 36158157; PMCID: PMC9494521.

7. Davenport A. Why is Intradialytic Hypotension the Commonest Complication of Outpatient Dialysis Treatments? Kidney Int Rep. 2022 Nov 10;8(3):405–418. doi: 10.1016/j.ekir.2022.10.031. PMID: 36938081; PMCID: PMC10014354.

8. Feng YM, Yang Y, Han XL, Zhang F, Wan D, Guo R. The effect of early versus late initiation of renal replacement therapy in patients with acute kidney injury: A meta-analysis with trial sequential analysis of randomized controlled trials. PLoS One. 2017 Mar 22;12(3):e0174158. doi: 10.1371/journal.pone.0174158. PMID: 28329026; PMCID: PMC5362192.

9. Grolleau F, Petit F, Gaudry S, Diard É, Quenot JP, Dreyfuss D, Tran VT, Porcher R. Personalizing renal replacement therapy initiation in the intensive care unit: a reinforcement learning-based strategy with external validation on the AKIKI randomized controlled trials. J Am Med Inform Assoc. 2024 Apr 19;31(5):1074–1083. doi: 10.1093/jamia/ocae004. PMID: 38452293; PMCID: PMC11031229.

10. Barbar SD, Binquet C, Monchi M, Bruyère R, Quenot JP. Impact on mortality of the timing of renal replacement therapy in patients with severe acute kidney injury in septic shock: the IDEAL-ICU study (initiation of dialysis early versus delayed in the intensive care unit): study protocol for a randomized controlled trial. Trials. 2014 Jul 7;15:270. doi: 10.1186/1745-6215-15-270. PMID: 24998258; PMCID: PMC4226975.

11. Metelli S, Chaimani A. Challenges in meta-analyses with observational studies. Evid Based Ment Health. 2020 May;23(2):83–87. doi: 10.1136/ebmental-2019-300129. Epub 2020 Mar 5. PMID: 32139442; PMCID: PMC10231593.

12. Xiao L, Jia L, Li R, Zhang Y, Ji H, Faramand A. Early versus late initiation of renal replacement therapy for acute kidney injury in critically ill patients: A systematic review and meta-analysis. PLoS One. 2019 Oct 24;14(10):e0223493. doi: 10.1371/journal.pone.0223493. PMID: 31647828; PMCID: PMC6812871.

13. Li X, Liu C, Mao Z, Li Q, Zhou F. Timing of renal replacement therapy initiation for acute kidney injury in critically ill patients: a systematic review of randomized clinical trials with meta-analysis and trial sequential analysis. Crit Care. 2021 Jan 6;25(1):15. doi: 10.1186/s13054-020-03451-y. PMID: 33407756; PMCID: PMC7789484.

14. Page MJ, McKenzie JE, Bossuyt PM, Boutron I, Hoffmann TC, Mulrow CD, Shamseer L, Tetzlaff JM, Akl EA, Brennan SE, Chou R, Glanville J, Grimshaw JM, Hróbjartsson A, Lalu MM, Li T, Loder EW, Mayo-Wilson E, McDonald S, McGuinness LA, Stewart LA, Thomas J, Tricco AC, Welch VA, Whiting P, Moher D. The PRISMA 2020 statement: an updated guideline for reporting systematic reviews. BMJ. 2021 Mar 29;372:n71. doi: 10.1136/bmj.n71. PMID: 33782057; PMCID: PMC8005924.

15. An N, Chen R, Bai Y, Xu M. Efficacy and prognosis of continuous renal replacement therapy at different times in the treatment of patients with sepsis-induced acute kidney injury. Am J Transl Res. 2021 Jun 15;13(6):7124–7131. PMID: 34306472; PMCID: PMC8290701.

16. Bai L, Luo L, Gao W, Bu C, Huang J. Curative effects of early continuous renal replacement therapy in cardiac failure combined with acute kidney injury. Int J Clin Exp Med. 2020;13(3):1612–1619.

17. STARRT-AKI Investigators; Canadian Critical Care Trials Group; Australian and New Zealand Intensive Care Society Clinical Trials Group; United Kingdom Critical Care Research Group; Canadian Nephrology Trials Network; Irish Critical Care Trials Group; Bagshaw SM, Wald R, Adhikari NKJ, Bellomo R, da Costa BR, Dreyfuss D, Du B, Gallagher MP, Gaudry S, Hoste EA, Lamontagne F, Joannidis M, Landoni G, Liu KD, McAuley DF, McGuinness SP, Neyra JA, Nichol AD, Ostermann M, Palevsky PM, Pettilä V, Quenot JP, Qiu H, Rochwerg B, Schneider AG, Smith OM, Thomé F, Thorpe KE, Vaara S, Weir M, Wang AY, Young P, Zarbock A. Timing of Initiation of Renal-Replacement Therapy in Acute Kidney Injury. N Engl J Med. 2020 Jul 16;383(3):240-251. doi: 10.1056/NEJMoa2000741. Erratum in: N Engl J Med. 2020 Jul 30;383(5):502. doi: 10.1056/NEJMx200016. PMID: 32668114.

18. Srisawat N, Laoveeravat P, Limphunudom P, Lumlertgul N, Peerapornratana S, Tiranathanagul K, Susantitaphong P, Praditpornsilpa K, Tungsanga K, Eiam-Ong S. The effect of early renal replacement therapy guided by plasma neutrophil gelatinase associated lipocalin on outcome of acute kidney injury: A feasibility study. J Crit Care. 2018 Feb;43:36–41. doi: 10.1016/j.jcrc.2017.08.029. Epub 2017 Aug 18. PMID: 28843662.

19 Lumlertgul N, Peerapornratana S, Trakarnvanich T, Pongsittisak W, Surasit K, Chuasuwan A, Tankee P, Tiranathanagul K, Praditpornsilpa K, Tungsanga K, Eiam-Ong S, Kellum JA, Srisawat N; FST Study Group. Early versus standard initiation of renal replacement therapy in furosemide stress test non-responsive acute kidney injury patients (the FST trial). Crit Care. 2018 Apr 19;22(1):101. doi: 10.1186/s13054-018-2021-1. PMID: 29673370; PMCID: PMC5909278.

20. Barbar SD, Clere-Jehl R, Bourredjem A, Hernu R, Montini F, Bruyère R, Lebert C, Bohé J, Badie J, Eraldi JP, Rigaud JP, Levy B, Siami S, Louis G, Bouadma L, Constantin JM, Mercier E, Klouche K, du Cheyron D, Piton G, Annane D, Jaber S, van der Linden T, Blasco G, Mira JP, Schwebel C, Chimot L, Guiot P, Nay MA, Meziani F, Helms J, Roger C, Louart B, Trusson R, Dargent A, Binquet C, Quenot JP; IDEAL-ICU Trial Investigators and the CRICS TRIGGERSEP Network. Timing of Renal-Replacement Therapy in Patients with Acute Kidney Injury and Sepsis. N Engl J Med. 2018 Oct 11;379(15):1431–1442. doi: 10.1056/NEJMoa1803213. PMID: 30304656.

21. Zarbock A, Kellum JA, Schmidt C, Van Aken H, Wempe C, Pavenstädt H, Boanta A, Gerß J, Meersch M. Effect of Early vs Delayed Initiation of Renal Replacement Therapy on Mortality in Critically Ill Patients With Acute Kidney Injury: The ELAIN Randomized Clinical Trial. JAMA. 2016 May 24-31;315(20):2190-9. doi: 10.1001/jama.2016.5828. PMID: 27209269.

22. Wald R, Adhikari NK, Smith OM, Weir MA, Pope K, Cohen A, Thorpe K, McIntyre L, Lamontagne F, Soth M, Herridge M, Lapinsky S, Clark E, Garg AX, Hiremath S, Klein D, Mazer CD, Richardson RM, Wilcox ME, Friedrich JO, Burns KE, Bagshaw SM; Canadian Critical Care Trials Group. Comparison of standard and accelerated initiation of renal replacement therapy in acute kidney injury. Kidney Int. 2015 Oct;88(4):897–904. doi: 10.1038/ki.2015.184. Epub 2015 Jul 8. PMID: 26154928.

23. Combes A, Bréchot N, Amour J, Cozic N, Lebreton G, Guidon C, Zogheib E, Thiranos JC, Rigal JC, Bastien O, Benhaoua H, Abry B, Ouattara A, Trouillet JL, Mallet A, Chastre J, Leprince P, Luyt CE. Early High-Volume Hemofiltration versus Standard Care for Post-Cardiac Surgery Shock. The HEROICS Study. Am J Respir Crit Care Med. 2015 Nov 15;192(10):1179–90. doi: 10.1164/rccm.201503-0516OC. PMID: 26167637.

24. Aklilu AM, Menez S, Baker ML, Brown D, Dircksen KK, Dunkley KA, Gaviria SC, Farrokh S, Faulkner SC, Jones C, Kadhim BA, Le D, Li F, Makhijani A, Martin M, Moledina DG, Coronel-Moreno C, O’Connor KD, Shelton K, Shvets K, Srialluri N, Tan JW, Testani JM, Corona-Villalobos CP, Yamamoto Y, Parikh CR, Wilson FP; KAT-AKI Team. Early, Individualized Recommendations for Hospitalized Patients With Acute Kidney Injury: A Randomized Clinical Trial. JAMA. 2024 Dec 24;332(24):2081–2090. doi: 10.1001/jama.2024.22718. PMID: 39454050; PMCID: PMC11669049.

25. Iyem H, Tavli M, Akcicek F, Büket S. Importance of early dialysis for acute renal failure after an open-heart surgery. Hemodial Int. 2009 Jan;13(1):55–61. doi: 10.1111/j.1542-4758.2009.00347.x. PMID: 19210279.

26. Gaudry S, Hajage D, Schortgen F, Martin-Lefevre L, Pons B, Boulet E, Boyer A, Chevrel G, Lerolle N, Carpentier D, de Prost N, Lautrette A, Bretagnol A, Mayaux J, Nseir S, Megarbane B, Thirion M, Forel JM, Maizel J, Yonis H, Markowicz P, Thiery G, Tubach F, Ricard JD, Dreyfuss D; AKIKI Study Group. Initiation Strategies for Renal-Replacement Therapy in the Intensive Care Unit. N Engl J Med. 2016 Jul 14;375(2):122–33. doi: 10.1056/NEJMoa1603017. Epub 2016 May 15. PMID: 27181456.

27. Bouman CS, Oudemans-Van Straaten HM, Tijssen JG, Zandstra DF, Kesecioglu J. Effects of early high-volume continuous venovenous hemofiltration on survival and recovery of renal function in intensive care patients with acute renal failure: a prospective, randomized trial. Crit Care Med. 2002 Oct;30(10):2205–11. doi: 10.1097/00003246-200210000-00005. PMID: 12394945.

28. Jiao R, Lu X, Liu M, Zhu J, Sun L, Liu N. Randomized Trial of Early vs Standard Renal Replacement Therapy in Patients With Acute Kidney Injury After Type A Aortic Dissection. Ann Thorac Surg. 2025 Jul;120(1):17–24. doi: 10.1016/j.athoracsur.2024.10.034. Epub 2025 Jan 4. PMID: 39761941.

29. Jamale TE, Hase NK, Kulkarni M, Pradeep KJ, Keskar V, Jawale S, Mahajan D. Earlier-start versus usual-start dialysis in patients with community-acquired acute kidney injury: a randomized controlled trial. Am J Kidney Dis. 2013 Dec;62(6):1116–21. doi: 10.1053/j.ajkd.2013.06.012. Epub 2013 Aug 8. PMID: 23932821.

30. Gaudry S, Hajage D, Martin-Lefevre L, Lebbah S, Louis G, Moschietto S, Titeca-Beauport D, Combe B, Pons B, de Prost N, Besset S, Combes A, Robine A, Beuzelin M, Badie J, Chevrel G, Bohé J, Coupez E, Chudeau N, Barbar S, Vinsonneau C, Forel JM, Thevenin D, Boulet E, Lakhal K, Aissaoui N, Grange S, Leone M, Lacave G, Nseir S, Poirson F, Mayaux J, Asehnoune K, Geri G, Klouche K, Thiery G, Argaud L, Rozec B, Cadoz C, Andreu P, Reignier J, Ricard JD, Quenot JP, Dreyfuss D. Comparison of two delayed strategies for renal replacement therapy initiation for severe acute kidney injury (AKIKI 2): a multicentre, open-label, randomised, controlled trial. Lancet. 2021 Apr 3;397(10281):1293–1300. doi: 10.1016/S0140-6736(21)00350-0. PMID: 33812488.

31. Jeong R, Bagshaw SM, Ghamarian E, Harvey A, Joannidis M, Kirkham B, McAuley D, Ostermann M, Quenot JP, Young PJ, Wald R; STandard vs. Accelerated initiation of Renal Replacement Therapy in Acute Kidney Injury (STARRT-AKI) Investigators. Time to Renal Replacement Therapy Initiation in Critically Ill Patients With Acute Kidney Injury: A Secondary Analysis of the Standard Versus Accelerated Initiation of Renal Replacement Therapy in Acute Kidney Injury (STARRT-AKI) Trial. Crit Care Med. 2025 Apr 1;53(4):e897–e907. doi: 10.1097/CCM.0000000000006616. Epub 2025 Mar 3. PMID: 40029115.

32. Jun M, Bellomo R, Cass A, Gallagher M, Lo S, Lee J; Randomized Evaluation of Normal Versus Augmented Level of Replacement Therapy (RENAL) Study Investigators. Timing of renal replacement therapy and patient outcomes in the randomized evaluation of normal versus augmented level of replacement therapy study. Crit Care Med. 2014 Aug;42(8):1756–65. doi: 10.1097/CCM.0000000000000343. PMID: 24717460.

33. Xia Y, Shi H, Wu W, Wang X. Effect of urinary NGAL on the timing of renal replacement therapy in patients with acute renal injury associated with sepsis. Med J Chin PLA. 2019;44(7):605–10

34. Ma H, Xu Y, Liu L. Effects of early continuous renal replacement therapy on treatment of sepsis patients complicated with acute kidney injury in stage 3. Chinese Journal of Nosocomiology 2020; 30: 2642–2646.

35. Sanghavi SF. Not So Fast: Kidney Replacement Therapy for Critically Ill Patients with AKI. Kidney360. 2022 May 11;3(7):1281-1284. doi: 10.34067/KID.0000932022. PMID: 35919533; PMCID: PMC9337890.

36. Moreira FT, Palomba H, Chaves RCF, Bouman C, Schultz MJ, Serpa Neto A. Early versus delayed initiation of renal replacement therapy for acute kidney injury: an updated systematic review, meta-analysis, meta-regression and trial sequential analysis of randomized controlled trials. Rev Bras Ter Intensiva. 2018 Jul-Sept;30(3):376-384. doi: 10.5935/0103-507X.20180054. PMID: 30328991; PMCID: PMC6180467.

37. Castro I, Relvas M, Gameiro J, Lopes JA, Monteiro-Soares M, Coentrão L. The impact of early versus late initiation of renal replacement therapy in critically ill patients with acute kidney injury on mortality and clinical outcomes: a meta-analysis. Clin Kidney J. 2022 May 12;15(10):1932–1945. doi: 10.1093/ckj/sfac139. PMID: 36158157; PMCID: PMC9494521.

