## supplementary file for "Early versus Delayed Initiation of Renal Replacement Therapy in Critically Ill Patients with Acute Kidney Injury: A Systematic Review and Meta-Analysis of Randomized Trials"

Supplementary Files

Search String

("acute kidney injury" OR AKI OR "acute renal failure") AND ("renal replacement therapy" OR RRT OR dialysis OR CRRT OR hemodialysis OR haemodialysis) AND (timing OR early OR delayed OR deferred OR accelerated OR initiation OR start)

Table S1. Summary Findings

| **Author Year** | **Country** | **Total** | **Male** | **Female** | **Criteria of Early RRT Initiation** | **Criteria for Delayed RRT Initiation** | **RRT Modality** | **RRT Initiation Time in Early** | **RRT Initiation time Delayed** | **SOFA Scores** | **SOFA Group** | **Age** | **Age Group** | **Early Number** | **Delayed Number** | **GRADE** | **Main Finding** |
| --- | --- | --- | --- | --- | --- | --- | --- | --- | --- | --- | --- | --- | --- | --- | --- | --- | --- |
| An et. Al. 2021 | China | 156 | 89 | 67 | CRRT immediately upon diagnosis of SAKI | CRRT only when disease progressed to acute indications or AKI stage 3 | CRRT | 1 | 6 | 22.14 ± 2.32 | >12 | 65.3 ± 10.3 | >65 | 78 | 78 | High | Early CRRT in sepsis-induced AKI improved renal function, reduced inflammatory markers, and lowered 60-day mortality compared with delayed CRRT |
| Bai et. Al. 2020 | China | 73 | 39 | 34 | Oliguria within 48 h, no hyperkalemia, Scr <200 µmol/L | ≥72 h persistent oliguria, hyperkalemia, Scr ≥200 µmol/L | CRRT | 48 | 72 | 20.18 ± 5.27 | >12 | **54.66 ± 8.55** | <65 | 37 | 36 | High | Early CRRT significantly improved hemodynamics, reduced Scr/BNP/CRP, shortened ICU and hospital stay, and lowered hospital mortality compared to delayed CRRT |
| Bagshaw et. Al. 2020 | Multicenter | 3019 | 1990 | 937 | Therapy started as soon as possible and **within 12 hours** of meeting eligibility | Initiated only if conventional indications developed (K⁺ ≥6 mmol/L, pH ≤7.20, HCO₃⁻ ≤12 mmol/L, severe respiratory failure with overload) **or if AKI persisted >72 hours** after randomization | CRRT | 6.1 | 31.1 | 11.6 ± 3.6 | <12 | 64.6 ± 14.3 | <65 | 1465 | 1462 | High | Accelerated RRT initiation **did not reduce 90-day mortality compared with a standard strategy**, but was associated with more RRT dependence and adverse events |
| Srisawat et. Al. 2018 | Thailand | 60 | 36 | 24 | Start CRRT **within 12 h of randomization** for patients with pNGAL ≥ 400 ng/mL | Start CRRT only if criteria met (severe acidosis, pulmonary edema, refractory hyperkalemia, anuria/oliguria, or BUN > 60 mg/dL | CRRT | 24 | 36 | 9.28 ± 3.62 | <12 | **66.8 ± 15.9** | >65 | 20 | 20 | High | Early RRT guided by pNGAL ≥ 400 ng/mL was feasible but did **not reduce 28-day mortality** compared to standard initiation |
| Lumlertgul et. Al. 2018 | Thailand | 162 | 58 | 60 | Within 6 hours after randomization | BUN ≥ 100 mg/dL, K+ > 6 mmol/L, HCO₃ < 12 mmol/L or pH < 7.15, PaO₂/FiO₂ < 200, or pulmonary edema | CVVH | 2 | 21 | 12.7 ± 3.3 | <12 | **67.5 ± 16.9** | >65 | 58 | 60 | High | Early RRT initiation guided by FST did not reduce mortality compared to standard initiation, but increased hypophosphatemia and catheter complications |
| Barbar et. Al. 2018 | France | 448 | 296 | 192 | Within 12 hours of documentation of RIFLE failure-stage AKI | After 48 hours if no spontaneous recovery and no emergency criteria (hyperkalemia >6.5 mmol/L, pH <7.15, or fluid overload) | CRRT | 7.6 | 51.5 | 12.2 ± 2.9 | >12 | 69.3 ± 11.6 | >65 | 246 | 242 | High | Early initiation of RRT in septic shock with severe AKI did not reduce 90-day mortality compared with delayed initiation |
| Zarbock et. Al. 2016 | Germany | 231 | 146 | 85 | Within 8 hours of diagnosis of KDIGO stage 2 AKI (2-fold creatinine rise or urine <0.5 mL/kg/h for ≥12 h, plus NGAL >150 ng/mL) | Within 12 h of stage 3 AKI (urine <0.3 mL/kg/h for ≥24 h, or 3-fold creatinine rise, or creatinine ≥4 mg/dL, or urgent indications: urea >100 mg/dL, K⁺ >6 mEq/L, anuria, refractory edema, etc.) | CVVH | 6 | 25.5 | 15.6 ± 2.3 | >12 | 65.7 ± 13.5 | >65 | 34 | 48 | High | Early RRT in critically ill AKI patients reduced 90-day mortality compared with delayed RRT |
| Canada et. Al. 2015 | Canada | 101 | 72 | 28 | Initiated within 12 h of meeting eligibility | RRT deferred unless urgent indications (K⁺ ≥6 mmol/L, bicarbonate <10 mmol/L, PaO₂/FiO₂ <200 + pulmonary edema) or AKI persisted >72 h | CRRT | 7.4 | 31.6 | 13.3 ± 2.5 | >12 | 62.2 ± 11.9 | <65 | 48 | 52 | High | Accelerated RRT initiation was feasible and safe but did not improve survival compared to standard initiation; about 25% of standard-arm patients recovered kidney function without RRT |
| Combes et. Al. 2015 | France | 224 | 117 | 47 | Immediate HVHF (80 ml/kg/h, max 8 L/h) within 3–24 h after cardiac surgery, maintained for 48 h | Only if persistent severe AKI (serum creatinine >354 µmol/L or 3× baseline, urine <0.3 ml/kg/h for 24 h, serum urea >36 mmol/L, or life-threatening hyperkalemia) | CVVH | 3 | 24 | 11.5 ± 2.8 | <12 | 61 ± 14 | <65 | 112 | 112 | High | Early HVHF after cardiac surgery shock did not reduce 30-day or longer-term mortality compared with delayed standard-volume CVVHDF |
| Akilu et. Al. 2024 | USA | 4003 | 2129 | 1874 | immediately | Start CRRT only if criteria met (severe acidosis, pulmonary edema, refractory hyperkalemia, anuria/oliguria, or BUN > 60 mg/dL | CRRT | 1 | 7 | 13.3 ± 2.5 | >12 | 72 ± 11 | >65 | 1999 | 2004 | High | Personalized kidney action team (KAT) recommendations increased implementation of diagnostic/medication changes but **did not reduce AKI progression, dialysis, or mortality compared with usual care** |
| Iyem et. Al. 2009 | Turkey | 185 | 117 | 68 | Dialysis initiated as soon as possible after ARF development | Dialysis started after 48 hours of ARF | CVVH | 1 | 48 | 12.4 ± 1.5 | >12 | 64.5 ± 5.2 | <65 | 5 | 6 | High | Early dialysis after cardiac surgery reduces morbidity (complications, ICU and hospital stay) but does not significantly reduce mortality |
| Gaudry et. Al. 2016 | France | 619 | 310 | 309 | Start **immediately after randomization**, within 6 hours of KDIGO stage 3 diagnosis | Start **only if** severe hyperkalemia, metabolic acidosis, pulmonary edema, BUN >112 mg/dl, or oliguria/anuria >72 hours occurred | CRRT | 2 | 57 | 10.9 ± 3.2 | <12 | 64.8 ± 14.2 | <65 | 129 | 134 | High | Delaying RRT avoided unnecessary dialysis in nearly half of patients without increasing mortality |
| Bouman et. Al. 2002 | Netherlands | 106 | 63 | 43 | Within 12 hrs of fulfilling inclusion criteria (oliguria ≤30 mL/hr for ≥6 hrs, despite resuscitation, inotropes, and high-dose diuretics) | Started when conventional indications were met: plasma urea ≥40 mmol/L, potassium ≥6.5 mmol/L, or severe pulmonary edema | CVVH | 6 | 42 | 10.3 ± 2.8 | <12 | 68 ± 13 | >65 | 70 | 36 | High | Early initiation or high-volume CVVH did not improve survival or renal recovery compared to delayed initiation or low-volume CVVH in critically ill patients with oliguric ARF |
| Jiao et. Al. 2025 | China | 212 | 141 | 71 | Within 6 hours of diagnosis of KDIGO stage 2 AKI | Within 8 hours of KDIGO stage 3 AKI or if absolute indications (urea >40 mmol/L, K+ >6 mmol/L despite therapy, pH <7.15) | CVVH | 6 | 36.3 | 14.6 ± 2.8 | <12 | 52.7 ± 10.9 | <65 | 106 | 106 | High | Early initiation of RRT within 6 hours of KDIGO stage 2 significantly reduced 90-day mortality compared to standard/delayed initiation |
| Jamale et. Al. 2013 | India | 208 | 141 | 67 | Dialysis if serum urea nitrogen >70 mg/dL and/or creatinine >7 mg/dL, irrespective of complications | Dialysis only when complications occurred (treatment-refractory hyperkalemia, volume overload, acidosis, uremic symptoms such as anorexia, nausea, or pericarditis) | Intermittent Hemodialysis | 7 | 48 | 7.6 ± 3.3 | <12 | 42.4 ± 15 | <65 | 102 | 106 | High | Earlier initiation of dialysis in community-acquired AKI did not improve survival and was associated with delayed kidney recovery |
| Gaudry et. Al. 2021 | France | 278 | 205 | 73 | RRT started **within 12 h after randomisation** once criteria (oliguria >72 h or BUN >112 mg/dL) were met | RRT postponed until **mandatory indication** (severe hyperkalaemia, metabolic acidosis, pulmonary oedema) or **BUN ≥140 mg/dL** | Intermittent Hemodialysis | 3 | 33 | 12 ± 3 | >12 | 65.5 ± 12 | >65 | 137 | 141 | High | A more-delayed RRT strategy (BUN ≥140 mg/dL or urgent indication) did **not increase RRT-free days** and was associated with **higher 60-day mortality** compared to standard-delayed strategy |
| Jeong et. Al. 2025 | Canada | 1462 | 731 | 731 | Start RRT within **12 hours of meeting eligibility criteria** | RRT only if conventional indications present: Serum potassium ≥ 6.0 mmol/L | CRRT | 12.1 | 24.5 | 13.7 ± 2.4 | >12 | 65 ± 11 | >65 | 226 | 677 | High | *Prolonged deferral of RRT in critically ill patients with severe AKI (without urgent indications) was not associated with increased 90-day mortality but was associated with higher RRT dependence at 90 days* |
| Jun et. Al. 2014 | Australia | 439 | 282 | 157 | Commencement <7.1 hrs from AKI diagnosis (RIFLE-I criteria) | Commencement ≥46 hrs from AKI diagnosis | CRRT | 7.1 | 46 | 2.0 ± 0.6 | <12 | 65.5 ± 13.2 | >65 | 39 | 44 | High | Earlier initiation of CRRT relative to RIFLE-I AKI onset did not significantly reduce 28- or 90-day mortality |
| Xia et. Al. 2019 | China | 60 | 33 | 27 | Sepsis and urinary, NGAL ≥ 1310 ng/mL | Serum potassium > 6.5 mmol/L;pH < 7.2; severe pulmonary edema | CRRT | 1 | 7 | 9.6 ± 3.2 | <12 | 66.4 ± 11.5 | >65 | 30 | 30 | High | Ealry Helped a lot in reducing the mortality |
| Ma et. Al. 2020 | China | 116 | 59 | 57 | Sepsis and urinary, NGAL ≥ 1310 ng/mL | Serum potassium > 6.5 mmol/L;pH < 7.2; severe pulmonary edema | CRRT | 58 | 58 | 12 ± 3 | >12 | 66.5 ± 7.82 | >65 | 58 | 58 | High | Ealry Helped a lot in reducing the mortality |

Risk of Bias

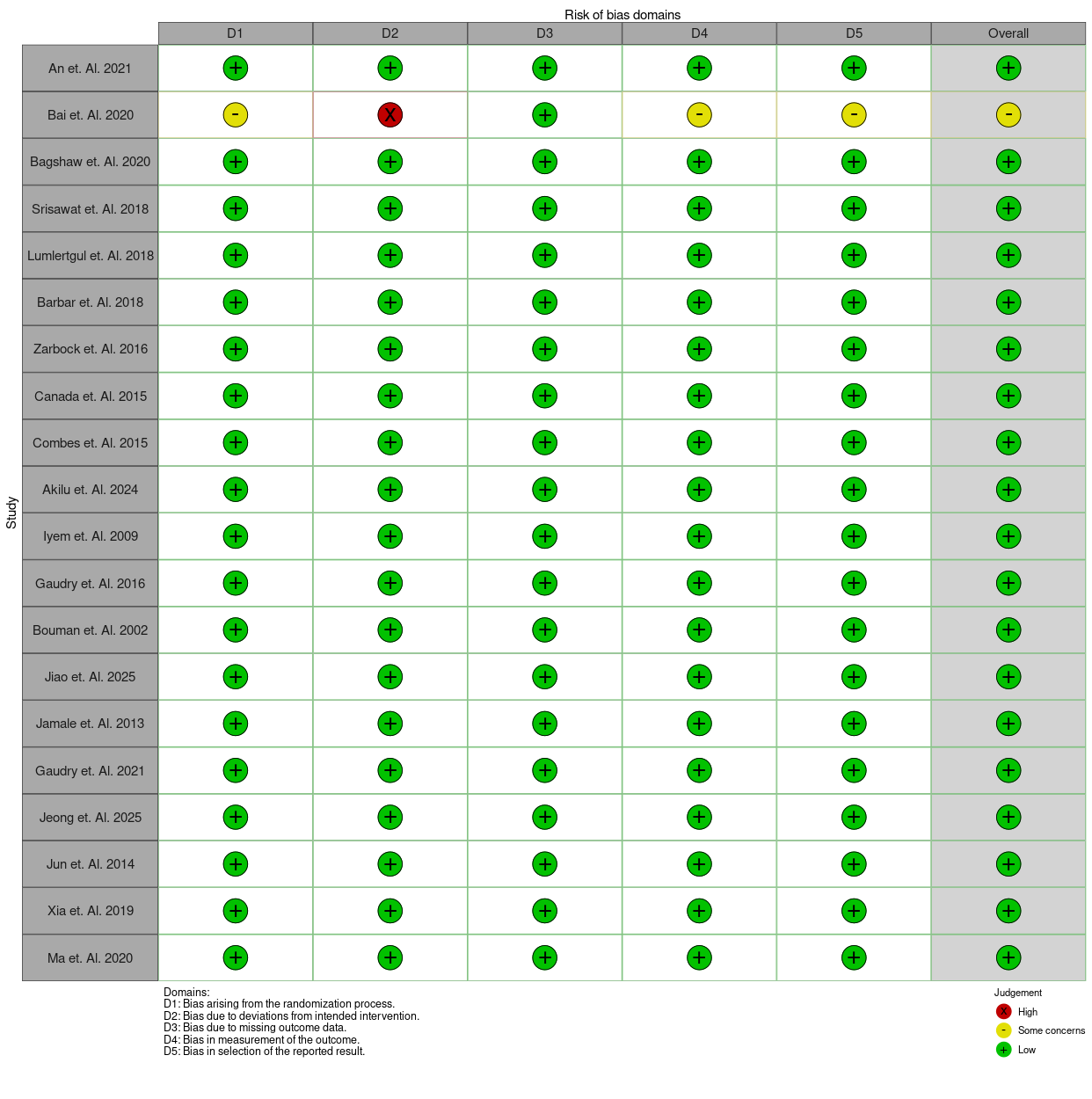
